# Long-Term Clinical and Sustained REMIssion in Severe Eosinophilic Asthma treated with Mepolizumab: The REMI-M study

**DOI:** 10.1101/2024.03.13.24304254

**Authors:** Claudia Crimi, Santi Nolasco, Alberto Noto, Angelantonio Maglio, Vitaliano Nicola Quaranta, Danilo Di Bona, Giulia Scioscia, Francesco Papia, Maria Filomena Caiaffa, Cecilia Calabrese, Maria D’Amato, Corrado Pelaia, Raffaele Campisi, Carolina Vitale, Luigi Ciampo, Silvano Dragonieri, Elena Minenna, Federica Massaro, Lorena Gallotti, Luigi Macchia, Massimo Triggiani, Nicola Scichilone, Giuseppe Valenti, Girolamo Pelaia, Maria Pia Foschino Barbaro, Giovanna Elisiana Carpagnano, Alessandro Vatrella, Nunzio Crimi, the Southern Italy Network on Severe Asthma Therapy

## Abstract

**Background:** Biological therapies, such as mepolizumab, have transformed the treatment of severe eosinophilic asthma. While mepolizumab’s short-term effectiveness is established, there is limited evidence on its ability to achieve long-term clinical remission.

**Objective:** To evaluate the long-term effectiveness and safety of mepolizumab, explore its potential to induce clinical and sustained remission, and identify baseline factors associated with the likelihood of achieving remission over 24 months.

**Methods:** The REMI-M is a retrospective, real-world, multicenter study that analyzed 303 severe eosinophilic asthma patients who received mepolizumab. Clinical, demographic, and safety data were collected at baseline, 3, 6, 12, and 24 months. The most commonly used definitions of clinical remission, which included no exacerbations, no oral corticosteroids (OCS) use, and good asthma control with or without assessment of lung function parameters, were adopted. Sustained remission was defined as reaching clinical remission at 12 months and maintaining it until the end of the 24-month period.

**Results:** Clinical remission rates ranged from 28.6% to 43.2% after 12 months and from 26.8% to 52.9% after 24 months, based on the different remission definitions. The proportion of patients achieving sustained remission varied between 14.6% to 29%. Factors associated with the likelihood of achieving clinical remission included the presence of aspirin-exacerbated respiratory disease, better lung function, male sex, absence of anxiety/depression, gastro-esophageal reflux disease, bronchiectasis, and reduced OCS consumption. Adverse events were infrequent.

**Conclusions:** This study demonstrates the real-world effectiveness of mepolizumab in achieving clinical remission and sustained remission in severe eosinophilic asthma over 24 months. The identification of distinct factors associated with the likelihood of achieving clinical remission emphasizes the importance of comprehensive management of comorbidities and timely identification of patients who may benefit from biologics.

**HIGHLIGHTS BOX:** *What is already known about this topic?:* Mepolizumab, an anti-IL-5 monoclonal antibody, has been shown to induce clinical remission after 12 months of treatment. However, long-term evidence remains limited.

*What does this article add to our knowledge?:* The REMI-M study investigated the effectiveness of mepolizumab in achieving clinical and sustained remission over 24 months.

*How does this study impact current management guidelines?:* Mepolizumab can elicit long-term clinical and sustained remission in a conspicuous proportion of patients with severe eosinophilic asthma, supporting its role as a possible disease-modifying agent. Management of comorbidities and timely identification of patients who may benefit from biological treatment are crucial for optimizing long-term outcomes.

## INTRODUCTION

Biological therapies have represented a significant advancement in the precision treatment of severe asthma. By targeting specific pathological drivers, these therapies potentially exhibit disease-modifying properties and have transformed the treatment of severe asthma (1–4). Mepolizumab is a humanized monoclonal antibody approved as add-on therapy for the treatment of severe eosinophilic asthma (SEA), and it neutralizes the circulating interleukin-5 (IL-5), which is involved in proliferation, differentiation, and survival of eosinophils (5).

The clinical effectiveness of mepolizumab has been evaluated in several randomized clinical trials, demonstrating a significant decrease in annual SEA exacerbations (6, 7) a reduction in systemic oral corticosteroids (OCS) intake (8) and improvements in symptom control and quality of life (9) with favorable long-term clinical effects and safety profile (10, 11). The benefits of mepolizumab on core asthma outcomes have been further confirmed in multiple real-world studies (12–15) that evaluated the overall effectiveness of mepolizumab in routine clinical practice, particularly in more complex patient populations, with multiple comorbidities that extend beyond those included in clinical trials (16–21).

These positive outcomes have led to a more ambitious treatment goal: the potential achievement of clinical remission of the disease. This represents a paradigm shift from a reactive ‘treat-to-failure’ approach to a proactive ‘treat-to-target’ strategy. Expert consensuses agreed on the concept that clinical remission is a multidimensional therapeutic goal which involves criteria aimed at achieving high level of disease control, including the absence of exacerbations, symptoms, OCS withdrawal and normalization/stabilization of lung function (22–24). However, the role and definition of clinical remission in SEA is still evolving (22, 25-27), while it is well-established in other clinical contexts, such as rheumatoid arthritis (28). Recently, several studies have evaluated the effectiveness of anti-IL-5 therapies in achieving clinical remission after one year of treatment (29). However, despite the growing body of evidence in this field, to date, studies assessing the impact of mepolizumab on achieving clinical remission at longer time points are lacking.

The REMI-M study aimed to assess the long-term effectiveness of mepolizumab in achieving clinical remission and sustained remission after one and two years of treatment.

## METHODS

### Study design

This retrospective observational multicenter study utilized anonymized medical records from patients referred to eleven specialized outpatients’ facilities participating in the “Southern Italy Network on Severe Asthma Therapy”. A complete list of all the participating centers is available in the Online Repository.

Data collection spanned from November 2019 to November 2022. This study adhered to the Declaration of Helsinki and received approval from the Ethics Committee “Catania 1” at the Policlinico University Hospital (Protocol Number 33/2020/PO). The institutional review boards at all participating centers approved the study protocol prior to the initiation of the study. All patients signed a written informed consent.

### Patient population

Data from adult patients (≥18 years) diagnosed with severe asthma according to the European Respiratory Society/American Thoracic Society (ERS/ATS) guidelines (30), which received add-on treatment with subcutaneous mepolizumab (100 mg, once every 4 weeks) were included. The indication for mepolizumab prescription followed the Italian eligibility criteria (baseline eosinophil count of ≥150 eosinophils/μL and a history of at least 300 eosinophils/μL in the previous 12 months, a minimum of 2 exacerbations in the previous year despite GINA step 5 maintenance therapy (31), and/or OCS maintenance treatment).

### Data collection

A shared database containing relevant variables for data acquisition was utilized across all participating centers. Demographic data and clinical variables were analyzed prior to the initiation of mepolizumab (baseline) and after 3, 6, 12 and 24 months of therapy. Clinical variables included blood eosinophil and basophils count, Asthma Control Test (ACT) (32) scores, fraction of exhaled nitric oxide (FeNO), pulmonary function tests, number of exacerbations, smoking habits and comorbidities such as anxiety/depression, gastroesophageal reflux disease (GERD), bronchiectasis, atopic dermatitis, aspirin-exacerbated respiratory disease (AERD), osteoporosis, and chronic rhinosinusitis with nasal polyps (CRSwNP).

Severe asthma exacerbations were defined as disease worsening requiring ≥3 days of treatment with systemic corticosteroids (or a doubling of the dose if already on OCS) (33). Exacerbations treated with <7 days apart were considered as the same episode.

Pulmonary function tests were performed in accordance with the ERS/ATS guidelines (34). Data on pre-bronchodilator forced expiratory volume in the first second (FEV_1_% and L), forced vital capacity (FVC%), FEV_1_/FVC%, and forced expiratory flow between 25% and 75% of FVC (FEF_25-75_%) were retrieved.

The fraction of exhaled nitric oxide (FeNO) was performed according to ERS/ATS recommendations (35).

Drug retention rate, defined as the percentage of patients remaining on mepolizumab over each time point.

Safety assessment involved reporting adverse events and monitoring laboratory values from the initial mepolizumab dose up to month 24. Their severity was defined in accordance with the World Health Organization guideline (36).

### Definitions of clinical remission and sustained remission

Currently, no single unified definition for clinical remission of SEA exists. Therefore, we evaluated clinical remission using different criteria:

i. *Three-component definition:* no annual exacerbations + no OCS + ACT ≥ 20 (37);
ii. *Four-component definition:* no annual exacerbations + no OCS + ACT ≥ 20 + lung function criteria (22–24);

As there are no widely accepted criteria to assess the normalization/stabilization of lung function, we performed a sensitivity analysis with different profiles of four-component definitions, including

the four most frequently lung function parameters considered by expert opinions (22–24). Thus, the different profiles of the *four-component definition* were classified as follows:

*Profile A:* no annual exacerbations + no OCS + ACT ≥ 20 + FEV_1_ ≥ 80%;
*Profile B:* no annual exacerbations + no OCS + ACT ≥ 20 + FEV_1_ +100mL from baseline;
*Profile C:* no annual exacerbations + no OCS + ACT ≥ 20 + FEV_1_ decline ≤ 5% from baseline;
*Profile D:* no annual exacerbations + no OCS + ACT ≥ 20 + FEV_1_ decline <100mL from the best value of the first 12 months (26).

Moreover, we explored the concept of *‘sustained remission’* defined as the obtainment of clinical remission at month 12 with retention of this status up to month 24.

### Statistical analysis

No formal sample size calculation was performed, and all available data on patients treated with mepolizumab from the “Southern Italy Network on Severe Asthma Therapy” were included. Data are presented as mean and standard deviation (±SD) for normally distributed continuous variables and as median and interquartile range (IQR) for continuous nonparametric variables. Categorical variables are expressed as numbers (n) and percentages (%). Analyses included only patients with available data at the given timepoint.

The normality of data distribution was checked using the Shapiro-Wilk and the Kolmogorov-Smirnov tests. Unpaired Student t-test or Mann-Whitney test were used for comparison of continuous variables at baseline. Fisher exact or McNemar tests were used for categorical variables. Mixed-effect model analysis, with Geisser-Greenhouse correction and Dunnett post hoc for repeated measures, was used to compare continuous outcomes from 3 to 24 months with baseline. Proportions of participants meeting the composite outcomes of clinical and sustained remission and individual remission criteria were reported descriptively.

Multiple logistic regression models were implemented to ascertain independent predictors of clinical remission and sustained remission. Variables with a *P* ≤.1 at univariate analysis were considered. Backward selection was applied, with an exclusion criterion of a *P* >.2. The variables were removed one at a time and the revised model was compared with the previous one using the Likelihood ratio test. The goodness of fit of the final models was confirmed by the Hosmer-Lemeshow test. Statistical analysis and figures were generated using Prism version 10.1.0 (GraphPad Software Inc., San Diego, California, USA) and SPSS Statistics 26 (IBM Corporation). A *P* <.05 (2-sided) was considered statistically significant.

## RESULTS

### Baseline patient demographic and clinical characteristics

Three hundred-three adult patients diagnosed with SEA and treated with mepolizumab were included in the study (Figure E1). Baseline demographic and clinical characteristics are outlined in Table 1. Overall, 63% were female, with a mean age of 56.9 ± 12.3 years and a median body mass index (BMI) of 27.1 ± 5.5 kg/m^2^. The median duration of the disease was 20 (10–30) years. Over one-third were current/ex-smokers. Regarding comorbidities, 14.9% were affected by anxiety/depression, 35% by gastro-esophageal reflux disease (GERD), 16.5% by bronchiectasis and 49.6% by chronic rhinosinusitis with nasal polyps (CRSwNP). All patients were prescribed high-dose inhaled corticosteroids and long-acting β-agonists, and 61.2% were also on long-acting muscarinic antagonists (LAMA); 69.4% of patients were on OCS, with a median maintenance dose of 10 mg/day (2.5-25) and 6.9% received prior treatments with anti-IgE (omalizumab).

**TABLE 1.**
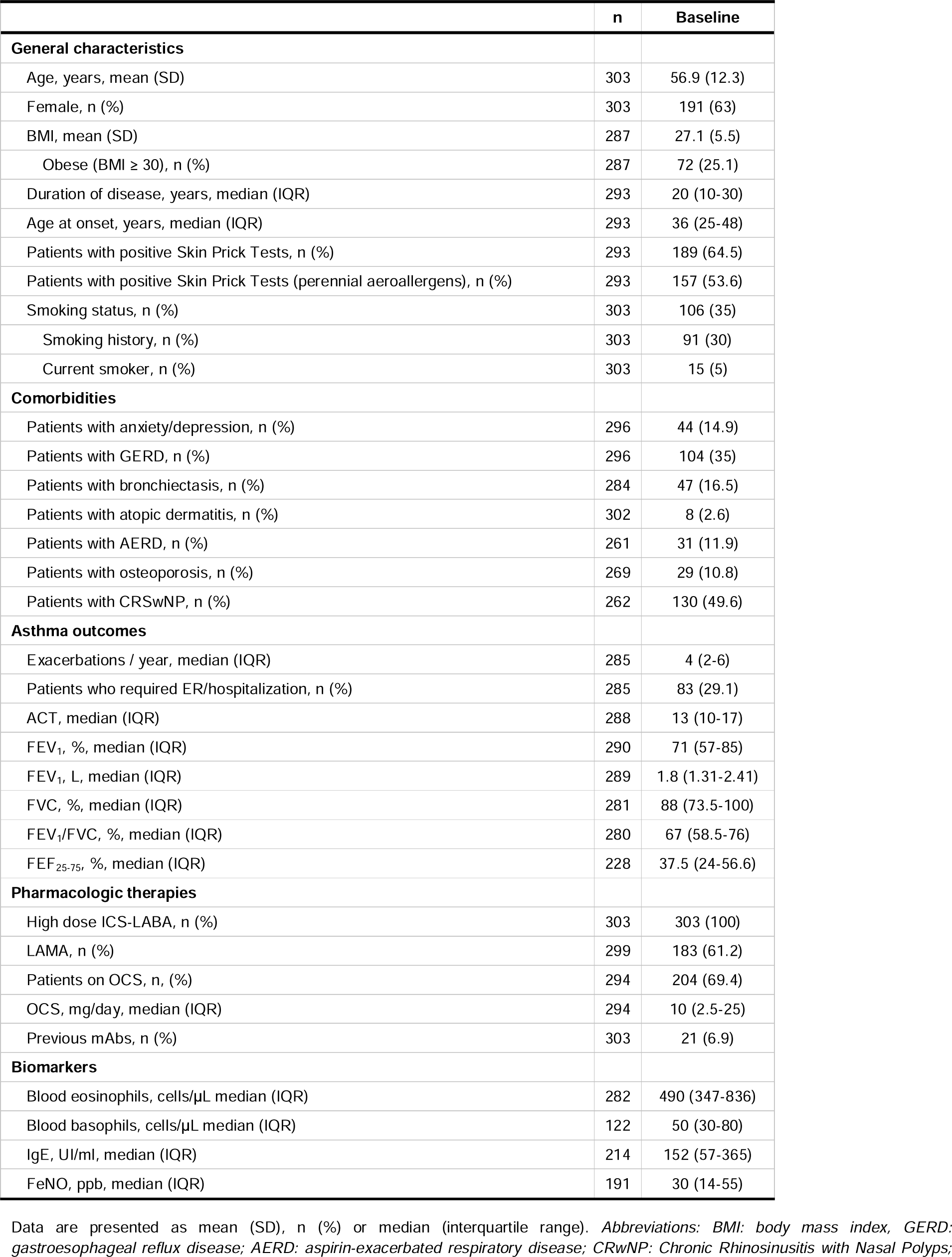
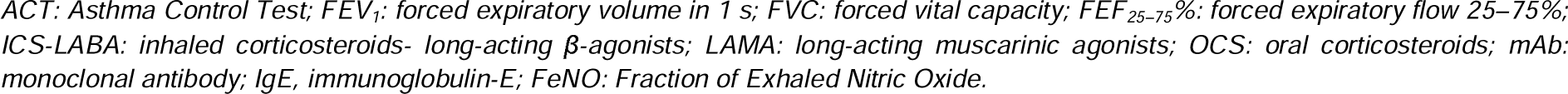
Patient characteristics at baseline.

|  | n | Baseline |
| --- | --- | --- |
| **General characteristics** |  |  |
| Age, years, mean (SD) | 303 | 56.9 (12.3) |
| Female, n (%) | 303 | 191 (63) |
| BMI, mean (SD) | 287 | 27.1 (5.5) |
| Obese (BMI $\geq 30$), n (%) | 287 | 72 (25.1) |
| Duration of disease, years, median (IQR) | 293 | 20 (10-30) |
| Age at onset, years, median (IQR) | 293 | 36 (25-48) |
| Patients with positive Skin Prick Tests, n (%) | 293 | 189 (64.5) |
| Patients with positive Skin Prick Tests (perennial aeroallergens), n (%) | 293 | 157 (53.6) |
| Smoking status, n (%) | 303 | 106 (35) |
| Smoking history, n (%) | 303 | 91 (30) |
| Current smoker, n (%) | 303 | 15 (5) |
| **Comorbidities** |  |  |
| Patients with anxiety/depression, n (%) | 296 | 44 (14.9) |
| Patients with GERD, n (%) | 296 | 104 (35) |
| Patients with bronchiectasis, n (%) | 284 | 47 (16.5) |
| Patients with atopic dermatitis, n (%) | 302 | 8 (2.6) |
| Patients with AERD, n (%) | 261 | 31 (11.9) |
| Patients with osteoporosis, n (%) | 269 | 29 (10.8) |
| Patients with CRSwNP, n (%) | 262 | 130 (49.6) |
| **Asthma outcomes** |  |  |
| Exacerbations / year, median (IQR) | 285 | 4 (2-6) |
| Patients who required ER/hospitalization, n (%) | 285 | 83 (29.1) |
| ACT, median (IQR) | 288 | 13 (10-17) |
| FEV<sub>1</sub>, %, median (IQR) | 290 | 71 (57-85) |
| FEV<sub>1</sub>, L, median (IQR) | 289 | 1.8 (1.31-2.41) |
| FVC, %, median (IQR) | 281 | 88 (73.5-100) |
| FEV<sub>1</sub>/FVC, %, median (IQR) | 280 | 67 (58.5-76) |
| FEF<sub>25-75</sub>, %, median (IQR) | 228 | 37.5 (24-56.6) |
| **Pharmacologic therapies** |  |  |
| High dose ICS-LABA, n (%) | 303 | 303 (100) |
| LAMA, n (%) | 299 | 183 (61.2) |
| Patients on OCS, n (%) | 294 | 204 (69.4) |
| OCS, mg/day, median (IQR) | 294 | 10 (2.5-25) |
| Previous mAbs, n (%) | 303 | 21 (6.9) |
| **Biomarkers** |  |  |
| Blood eosinophils, cells/μL median (IQR) | 282 | 490 (347-836) |
| Blood basophils, cells/μL median (IQR) | 122 | 50 (30-80) |
| IgE, UI/ml, median (IQR) | 214 | 152 (57-365) |
| FeNO, ppb, median (IQR) | 191 | 30 (14-55) |

Data are presented as mean (SD), n (%) or median (interquartile range). Abbreviations: BMI: body mass index, GERD: gastroesophageal reflux disease; AERD: aspirin-exacerbated respiratory disease; CRwNP: Chronic Rhinosinusitis with Nasal Polyps;
ACT: Asthma Control Test; FEV<sub>1</sub>: forced expiratory volume in 1 s; FVC: forced vital capacity; FEF<sub>25–75</sub>%; forced expiratory flow 25–75%; ICS-LABA: inhaled corticosteroids- long-acting β-agonists; LAMA: long-acting muscarinic agonists; OCS: oral corticosteroids; mAb: monoclonal antibody; IgE, immunoglobulin-E; FeNO: Fraction of Exhaled Nitric Oxide.

### Effectiveness of mepolizumab therapy

The annual exacerbation rate significantly decreased from 4 (2–6) exacerbation/year at baseline to 1 (0–2) after 12 months and to 0 (0–1) after 24 months (*P*< .0001) (Figure 1A). Indeed, the percentage of patients requiring emergency room access or hospitalization decreased from 29.1% to 2% after 24 months (Figure 1B). The ACT score significantly improved from 13 (IQR, 10-17) to 21 (IQR, 18-23) already after 3 months (*P*< .0001) and reached the median value of 22 (20–24) at month 24 (*P*< .0001) (Figure 1C). Significant improvements in lung function were found, with the increase in FEV_1_% (*P*< .0001) (Figure 1D), FEV_1_ (L), FVC%, FEV_1_/FVC and FEF_25-75_% (Table E1). The proportion of patients on OCS decreased from 69.4% at baseline to 16.5% after 24 months (*P*< .0001). The median daily OCS dose decreased from 10 mg/day (2.5-25) to 0.0 mg/day (0–0) at month 24 (*P*< .0001) (Figure 1E). The blood eosinophils (Figure 1F) and basophils sharply dropped during treatment (*P*< .0001 at each timepoint) (Table E1). No significant changes in FeNO were detected (Table E1) (*P*< .2416 at 24 months).

**Figure 1.**
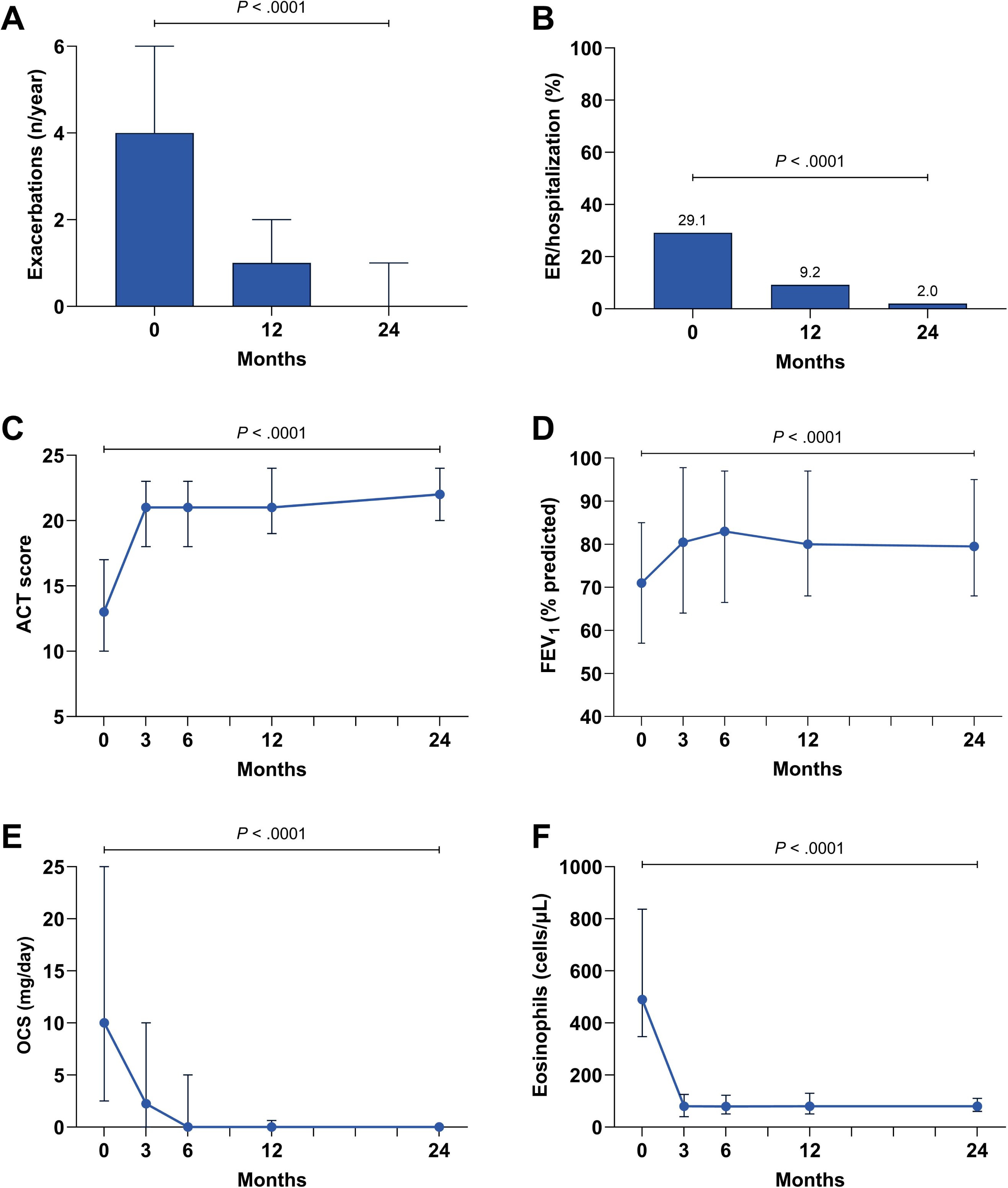
Effectiveness of mepolizumab on clinical outcomes. *Abbreviations: ACT, asthma control test; FEV_1_, forced expiratory volume in the 1st second; OCS, oral corticosteroids (prednisone equivalent dose)*.

### Proportion of patients who met individual criteria for the definitions of clinical remission

Figure 2 illustrates the percentage of patients who met each criterion used for the different definitions of clinical remission. Notably, about half of the cohort had no annual exacerbations (Figure 2A) at month 24. There was an increase in the number of patients who discontinued OCS, with 83.6% being OCS-free after 24 months (Figure 2B). The ACT score rapidly improved, with 64.4% of patients reaching the threshold score of 20 within just 3 months and 78% at 24 months (Figure 2C). The percentages of both the FEV_1_ ≥80% (Figure 2D) and the +100mL increase in FEV_1_ from baseline (Figure 2E) progressively improved up to 6 months, followed by a gradual decrease. A FEV_1_ decline ≤ 5% from baseline was reported in over 80% of the entire cohort throughout the study period (Figure 2F). However, a FEV_1_ decline <100mL from the best value of the first 12 months was obtained only by 49.6% at month 24 (Figure 2G).

**Figure 2.**
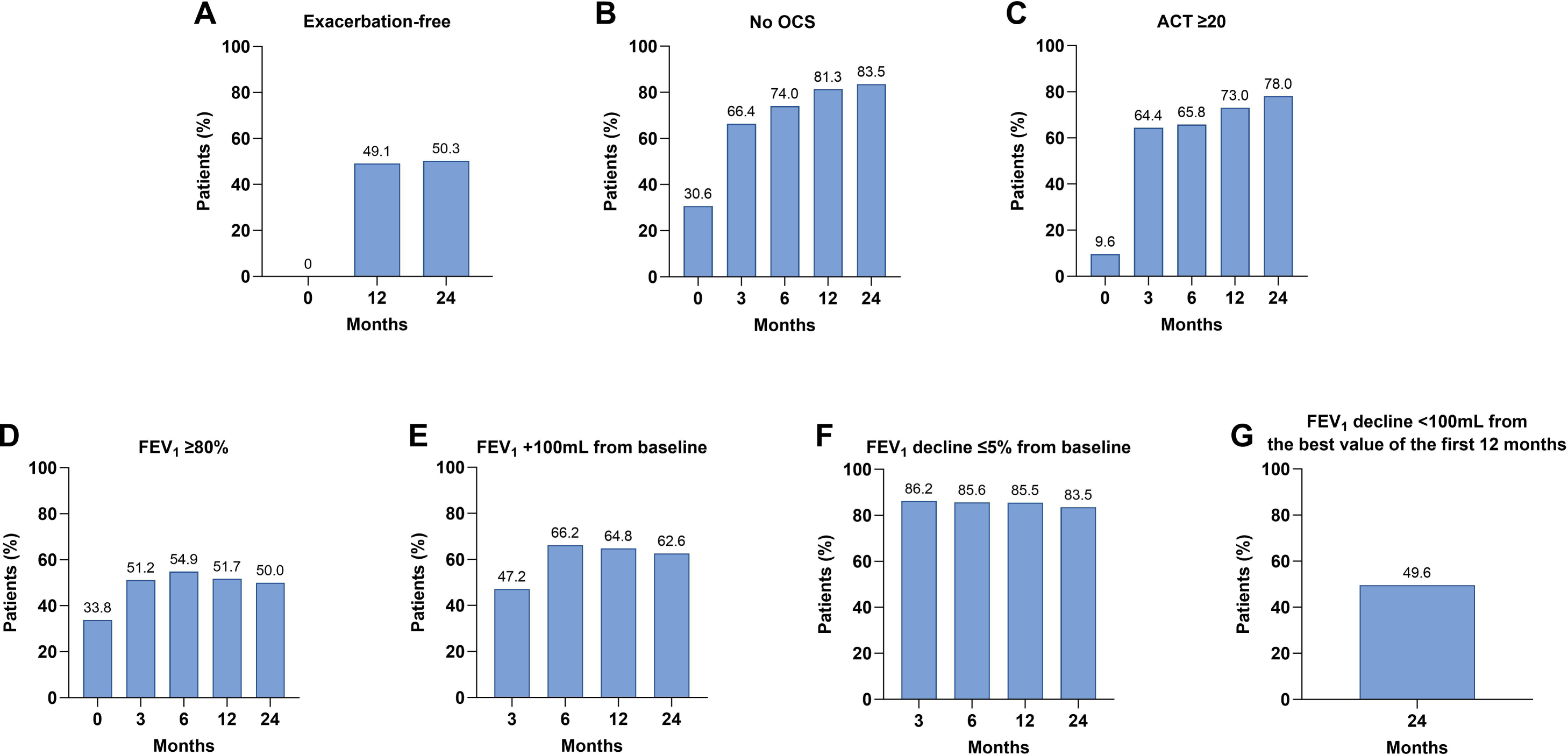
Percentages of patients who met the criteria used to define clinical remission. *Abbreviations: ACT, asthma control test; OCS, oral corticosteroids (prednisone equivalent dose), FEV_1_, forced expiratory volume in the 1st second*.

### Proportion of patients achieving long-term clinical and sustained remission

Two hundred eighteen patients completed 12 months of treatment, whereas 147 were treated for 24 months (Figure E1). The number of subjects with all the explanatory variables for each definition of remission is presented in Table E2 and Figure 4.

#### Three-component definition: no annual exacerbations + no OCS + ACT ≥ 20

Overall, 43.2% achieved all three main criteria after 12 months and 52.9% after 24 months (Figure 3); 29.7% remained in sustained remission during the observed period (Figure 3).

**Figure 3.**
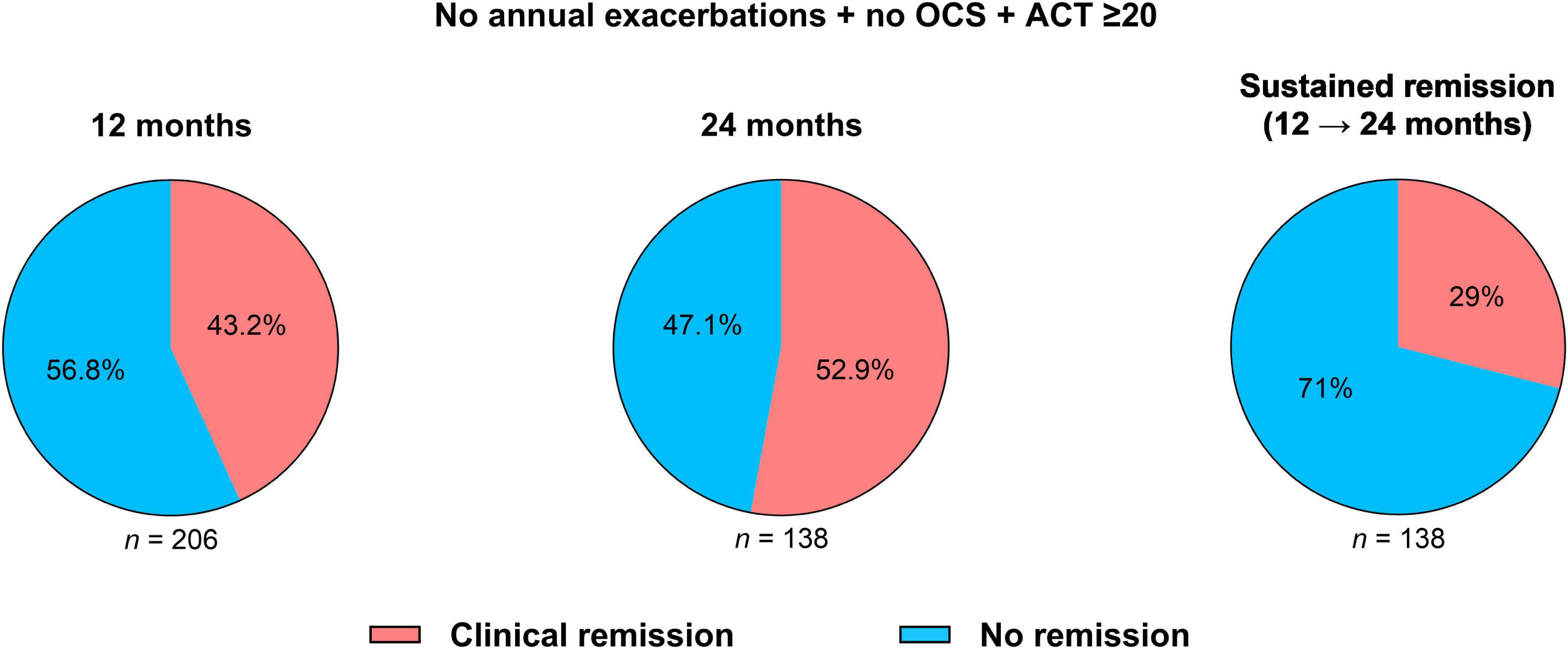
Percentages of patients achieving the three-component definition of clinical remission and sustained remission after 12 and 24 months of treatment. *Abbreviations: ACT, asthma control test; OCS, oral corticosteroids (prednisone equivalent dose)*.

#### Four-component definition (Profile A): no annual exacerbations + no OCS + ACT ≥ 20 + FEV_1 ≥_ 80%

31.3% and 39.1% of patients fulfilled the criteria at 12 and 24 months, but only 21.9% remained in sustained remission throughout the second year (Figure 4A).

**Figure 4.**
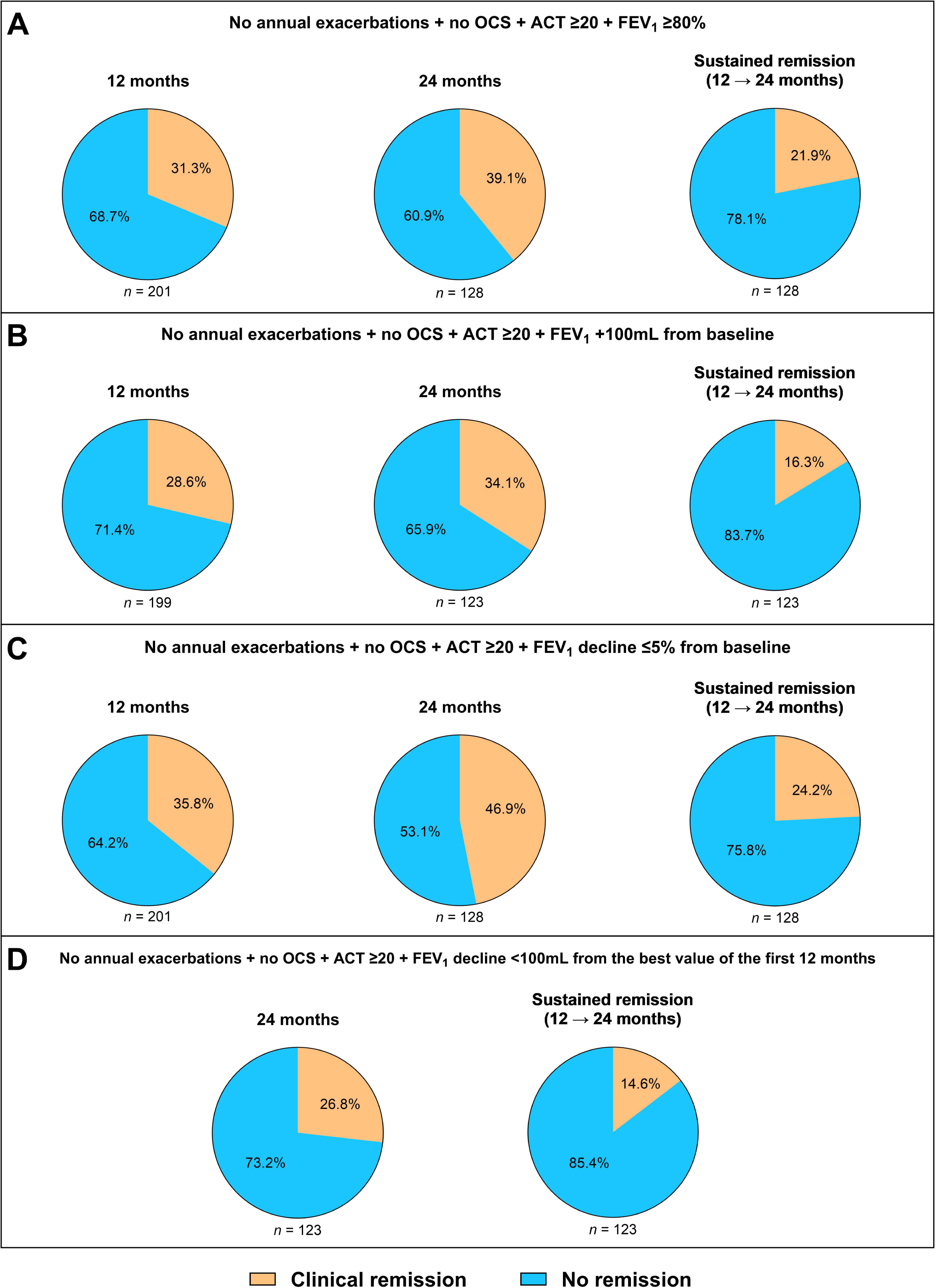
Percentages of patients achieving the four-component definitions of clinical remission and sustained remission after 12 and 24 months of treatment. *Abbreviations: ACT, asthma control test; OCS, oral corticosteroids (prednisone equivalent dose), FEV_1_, forced expiratory volume in the 1st second*.

#### Four-component definition (Profile B): no annual exacerbations + no OCS + ACT ≥ 20 + FEV_1_ +100mL from baseline

28.6% and 34.1% of patients were in clinical remission at months 12 and 24, respectively, with 16.3% achieving sustained remission (Figure 4B).

#### Four-component definition (Profile C): no annual exacerbations + no OCS + ACT ≥ 20 + FEV_1_ decline _≤_5% from baseline

35.8% and 46.9% of patients were in clinical remission after 12 and 24 months, respectively, and 24.2% reached sustained remission (Figure 4C).

#### Four-component definition (Profile D): no annual exacerbations + no OCS + ACT ≥ 20 + FEV1 decline <100mL from the best value of the first 12 months

Only 26.8% of patients were in clinical remission and 14.6% in sustained remission at month 24, respectively (Figure 4D).

### Baseline predictors of long-term clinical and sustained remission

According to the different definitions of remission, we found distinct characteristics of patients who achieved clinical remission after 24 months.

#### Three-component definition: no annual exacerbations + no OCS + ACT ≥ 20

In the univariate analysis, patients who reached clinical remission after 24 months were fewer smokers, had lower incidence of anxiety/depression and GERD and decreased usage of LAMA and OCS while exhibiting better lung function (Table E3). The regression model revealed that being female reduced the likelihood of clinical remission by 79.6% (*P*= .0030), with current/ex-smokers having a 73.1% reduction in odds (*P*= .0110). However, each unit increase in FEV_1_/FVC was associated with a 4.1% increased probability of achieving remission (*P*= .0200) (Figure 5). Patients who achieved sustained remission had less anxiety/depression, reduced OCS use and better lung function (Table E4). However, in the multivariate model, only OCS intake was retained; specifically, each mg increase in the daily OCS dosage was linked to a 7% decrease in the probability of achieving sustained remission (*P*= .00010) (Figure 5).

**Figure 5.**
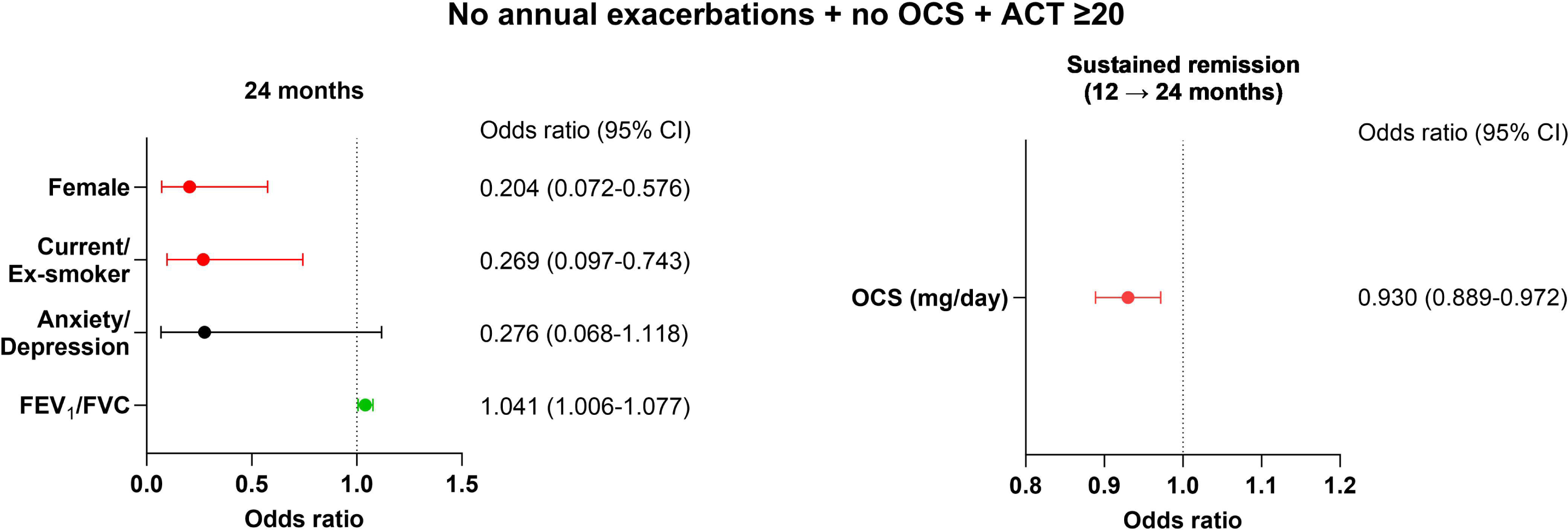
Baseline factors associated with the long-term three-component definition of clinical and sustained remission. Variables presented in green favour remission, whereas red ones exert a negative effect. *Abbreviations: OCS, oral corticosteroids (prednisone equivalent dose), FEV_1_, forced expiratory volume in the 1st second; FVC: forced vital capacity*.

#### Four-component definition (Profile A): no annual exacerbations + no OCS + ACT ≥ 20 + FEV_1_ ≥ 80%

In the univariate analysis of baseline characteristics (Table E5), the group of patients who achieved clinical remission after 24 months had a lower prevalence of smokers and less anxiety/depression but more aspirin-exacerbated respiratory disease (AERD). They also had better ACT scores and lung function and were less likely to be on LAMA and OCS therapy. In the regression model, the likelihood of achieving clinical remission increased by 5-fold in patients with AERD (*P*= .0070), by 92.5% in those with anxiety/depression (*P*= .0130) and by 2.9% for every unit increase in FEF_25-75_% (*P*= .0050) (Figure 6A). Those who achieved sustained remission had less frequently bronchiectasis, showed a more preserved lung function, were less likely to be on LAMA and OCS and had higher FeNO at baseline (Table E6). In the regression model, each unit increase in FEV_1_% and FEF_25-75_% improved the odds of sustained remission by 13.7% (*P*= .0410) and 3.6% (*P*= .0010), respectively (Figure 6A).

**Figure 6.**
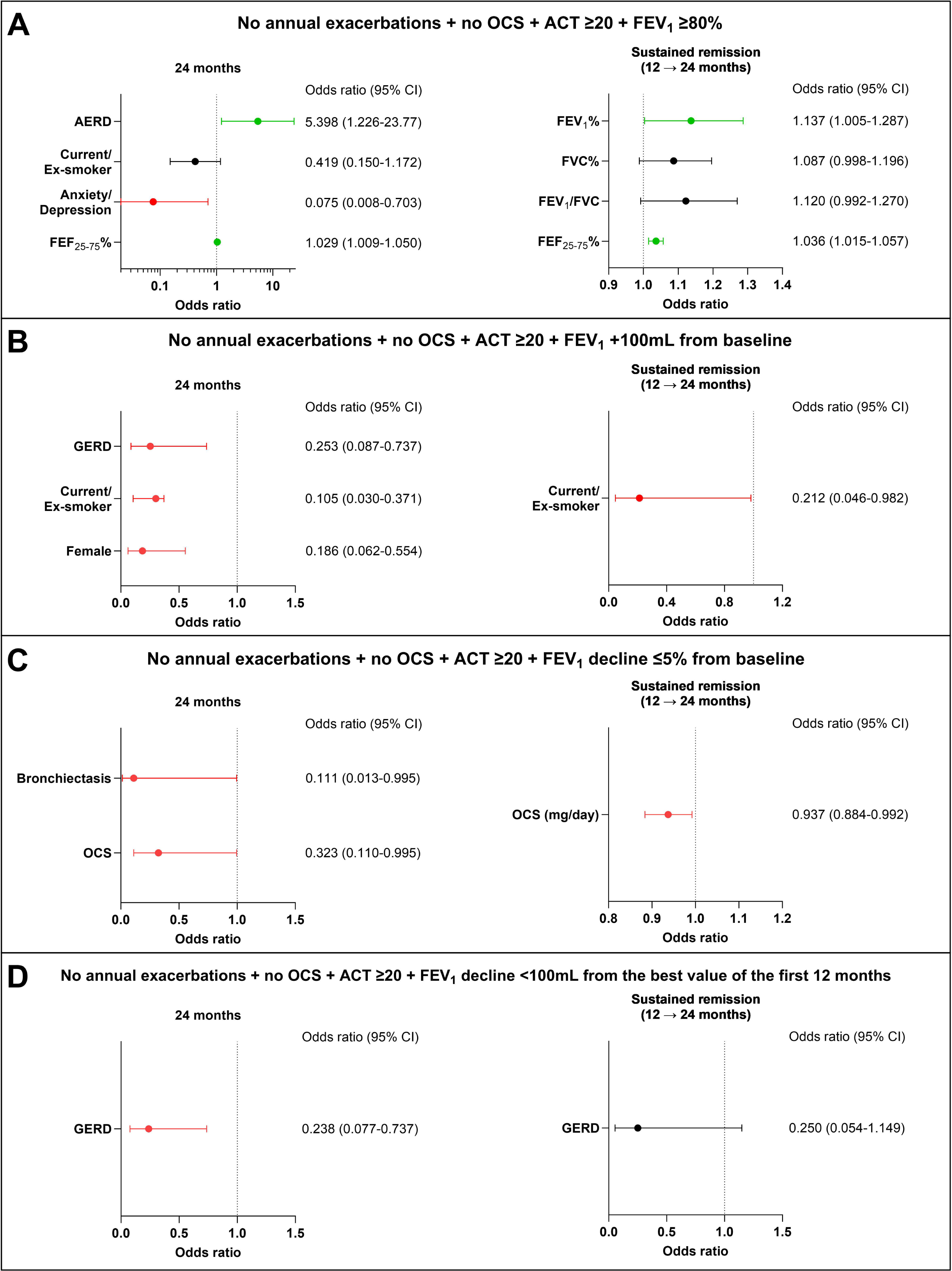
Baseline factors associated with the long-term four-component definitions of clinical and sustained remission. Variables presented in green favour remission, whereas those in red exert a negative effect. *Abbreviations: OCS, oral corticosteroids (prednisone equivalent dose), FEV_1_, forced expiratory volume in the 1st second, AERD: aspirin-exacerbated respiratory disease; GERD: gastroesophageal reflux disease; FVC: forced vital capacity; FEF_25–75_%: forced expiratory flow 25– 75%*.

#### Four-component definition (Profile B): no annual exacerbations + no OCS + ACT ≥ 20 + FEV_1_ +100mL from baseline

Female gender, smoking, anxiety/depression, and GERD were more frequent in patients not achieving clinical remission at month 24 (Table E7). Multivariate analysis confirmed that the odds of achieving clinical remission decreased by 74.7% in patients with GERD (*P*= .0030), 89.5% in current/ex-smokers (*P*< .0001) and 81.4% for females (*P*= .0120), respectively (Figure 6B). The sole variable retained from univariate analysis (Table E8) for sustained remission was smoking status (*P*= .0470) (Figure 6B).

#### Four-component definition (Profile C): no annual exacerbations + no OCS + ACT ≥ 20 + FEV_1_ decline _≤_5% from baseline

Female gender, smoking, anxiety/depression, GERD, bronchiectasis, LAMA, and OCS use were more prevalent among individuals who did not reach clinical remission at month 24 (Table E9). In the regression analysis, bronchiectasis (*P*= .0450) and OCS use (*P*= .0410) reduced the probability of clinical remission by 89.9% and 67.7%, respectively (Figure 6C). A lower dose of OCS and greater baseline blood basophils and FeNO levels were associated with sustained remission (Table E10). Each mg increase in the daily OCS dose decreased the likelihood of achieving sustained remission by 6.3% (*P*= .0260) (Figure 6C).

#### Four-component definition (Profile D): no annual exacerbations + no OCS + ACT ≥ 20 + FEV_1_ decline <100mL from the best value of the first 12 months

Anxiety/depression and GERD were more frequent in patients who had not achieved clinical remission by month 24 (Table E11). In regression analysis, GERD had a negative impact, lowering the likelihood of clinical remission by 76.2% (*P*= .0130) (Figure 6D). A lower prevalence of bronchiectasis and higher FeNO levels were predictors of sustained remission at univariate analysis (Table E12); however, no variables were retained in the regression model (Figure 6D).

### Retention rate

At the 12-month follow-up, mepolizumab retention rate was 94%. Fourteen patients (6%) were switched to different biologics (Table E13). At the 24-month follow-up, 147 out of 166 (88.6%) patients were still on-treatment with mepolizumab (Table E13).

### Safety

Table 2 shows the number and type of adverse events that occurred during the study period. Only one serious adverse event was reported. None of the patients discontinued mepolizumab due to adverse events.

**TABLE 2.** Adverse events.

|  | n | 12 months | n | 24 months |
| --- | --- | --- | --- | --- |
| **Total adverse event, n (%)** | 218 | 7 (3.2) | 147 | 1 (0.7) |
| **Mild-moderate adverse event, n (%)** | 218 | 6 (2.8) | 147 | 1 (0.7) |
| Fever, n (%) | 218 | 3 (1.4) | 147 | 0 (0) |
| Urticaria, n (%) | 218 | 2 (0.9) | 147 | 0 (0) |
| Otitis, n (%) | 218 | 1 (0.5) | 147 | 0 (0) |
| Atrial fibrillation, n (%) | 218 | 0 (0) | 147 | 1 (0.7) |
| **Serious adverse event, n (%)** | 218 | 1 (0.5) | 147 | 0 (0) |
| Bronchospasm, n (%) | 218 | 1 (0.5) | 147 | 0 (0) |
| **Adverse event requiring treatment discontinuation, n (%)** | 218 | 0 (0) | 147 | 0 (0) |

## DISCUSSION

In this large, multicenter, real-world, cohort study, treatment with mepolizumab was associated with an overall clinical remission rate after 12 months ranging from 28.6% to 43.2% and after 24 months obtained by 26.8 to 52.9% of patients, depending on the definition applied, with one in five patients able to achieve sustained remission from months 12 to 24. To the best of our knowledge, this is the first study to report real-world data on clinical and sustained remission after 24 months of mepolizumab treatment in a large cohort of SEA patients.

In our heterogeneous population affected by a wide range of comorbidities, the overall effectiveness of mepolizumab surpassed those reported in clinical trials (6–9) while it was comparable or superior in some cases to the limited number of long-term (24 months or more) real-world studies conducted to date (14, 15, 38-41). Remarkably, we observed a 75% reduction in the exacerbation rate after 12 months of treatment, with a 100% decrease at month 24. Interestingly, the reduction in exacerbation rates was more pronounced compared to the MENSA (7) (−53%) and MUSCA (9) (−58%) trials but similar to those reported in real-life studies such as REALITI-A (−79%) (39) and Matucci et al. (−84%) (15). These findings are valuable from both physicians’ and patients’ perspectives, as the frequency and severity of exacerbations are associated with accelerated lung function decline (42) and worse quality of life (6, 15, 43).

Another important finding of our study pertains the use of OCS. Indeed, more than 80% of patients were OCS-free at 12 months and 24 months, respectively. These findings are consistent with data reported in REDES (29) (81% of patients were OCS-free at 1 year) and by Bagnasco et al. (14) (79% of patients were OCS-free at 1 year and 89% at 2 years, respectively,) but superior to REALITI-A (41) (57% of patients were OCS-free at 2 years).

In terms of clinical remission, to date, only a few studies have investigated the effects of mepolizumab, with two studies assessing its effectiveness after one year of treatment (29, 44), one study evaluating the super-response to anti-IL-5/Rα biologics after 2 years (45).

The definitions of remission may play a crucial role in the treat-to-target strategy in SEA; however, currently, there are no universal definition or criteria to define clinical remission in SEA. In REMI-M, we explored a three-component model for defining clinical remission that intentionally excluded lung function measurements in favor of an overall disease assessment, including resolution of symptoms, absence of systemic steroids use and no interference with patients’ activities of daily living, which we considered essential aspects from the patients’ perspective. We also explored the most used composite criteria for defining clinical and sustained remission from a research/clinician point of view to ensure adequate reproducibility and allow for direct comparison with prior studies (26). Specifically, according to the lung function parameter adopted, the percentage of patients achieving clinical remission at 12 months ranged from 28.6% to 35.8%, in concordance with the already available evidence (29, 37, 44). However, the clinical remission rate after one year in our study was higher compared to the Danish (19%) and UK (18.3%) national registry cohorts (46, 47). This difference may be attributed to the centers involved in the management of SEA patients on biologics; thus, the uniform and systematic clinical approach adopted by the dedicated severe asthma facilities within the “Southern Italy Network on Severe Asthma Therapy” might have contributed positively to our findings, in contrast with national registries, which could be subject to variations in clinical practice. Furthermore, by focusing solely on mepolizumab, our study did not account for the potential impact of anti-IgE treatments on the overall clinical remission rates. Notably, anti-IgE therapy has demonstrated lower remission rates than anti-IL-5, likely due to the IgE’s role as a downstream mediator of inflammation, resulting in less pronounced effects when targeted (48).

Significantly, clinical remission rates increased over the 24-month period, escalating from 26.8% to 52.9%. This increase may be attributed to a concomitant progressive reduction in OCS use and the rise in the proportion of subjects with ACT ≥20.

Given that anti-IL-5 therapy is known to mitigate the decline in lung function (49), we have introduced, for the first time, an assessment of lung function decline from the best value obtained within the first 12 months of treatment. The accelerated lung function decline among SEA patients reflects uncontrolled inflammatory processes and airway remodelling. Consequently, assessing this decline over the long-term could bridge the gap between the concept of clinical and biological remission (26). Indeed, the adoption of this stringent parameter revealed that less than half of the patients achieved it, substantially influencing both clinical and sustained remission rates.

Sustained remission rates were significantly lower, ranging from 14.6% to 29%; this was primarily attributed to exacerbations that occurred during the second year of treatment, along with a slight decline in lung function observed between months 6 to 24.

Our study reveals that patients who achieved long-term clinical and sustained remission exhibit distinctive baseline characteristics. Overall, non-remission was commonly associated with female sex, coexisting anxiety or depression, GERD, bronchiectasis, previous or active smoking, and OCS use. In contrast, remission was more likely in patients with AERD and a better baseline lung function. Notably, these determinants concur with findings from previous studies on clinical remission after 12 months (37, 44, 46, 47), thus confirming their influence also in the long-term. These findings highlight the crucial role that effective management of comorbidities plays in the treatment of SEA and suggest that the timely introduction of biologics may improve the likelihood of achieving clinical remission, emphasizing the importance of prompt patients’ referral to SEA dedicated facilities. We believe that our data have important clinical implications in the management of SEA patients and provide useful insights for future research.

Our study has several strengths. First, it included a large and heterogeneous population of SEA patients. Indeed, unlike clinical trials and as in every real-world evidence, the study was not constrained by stringent patient selection based on specific inclusion criteria, encompassing a broader patient population that reflects the complexities and diversity of everyday clinical practice. Second, the follow-up period of 24 months allowed for long-term assessment. However, the study also has limitations, including its retrospective design and its related potential variability in reporting. Nonetheless, the data collection protocol was rigorously followed across multiple sites within a network with recognized expertise and extensive experience in SEA management and biological treatment, ensuring consistency in patient criteria for SEA assessment and mepolizumab prescription. Finally, as is common with retrospective study designs, there is a possibility that adverse events were underreported.

In summary, our study provides valuable real-world evidence confirming the effectiveness of mepolizumab in achieving clinical and sustained remission in SEA patients over 12 and 24 months, identifying factors associated with the likelihood of achieving clinical remission.

## Supporting information

Online Repository

## Data Availability

All data produced in the present study are available upon reasonable request to the authors

## ABBREVIATIONS

ACT: Asthma Control Test
AERD: Aspirin-exacerbated respiratory disease
ERS/ATS: European Respiratory Society/American Thoracic Society
CRSwNP: Chronic rhinosinusitis with nasal polyps
OCS: Oral corticosteroids (prednisone)
SEA: Severe eosinophilic asthma
FEF_25-75_%: Forced expiratory flow between 25% and 75% of FVC
FeNO: Fractional exhaled nitric oxide
FEV_1_: Forced expiratory volume in the 1st second
FVC: Forced vital capacity
GERD: Gastroesophageal reflux disease
LAMA: Long-acting muscarinic antagonists

