## Supplementary material for "Long-Term Clinical and Sustained REMIssion in Severe Eosinophilic Asthma treated with Mepolizumab: The REMI-M study": Online Repository

***List of centers***

1. Respiratory Medicine Unit - Policlinico “G. Rodolico-San Marco” University Hospital, Catania;
2. Pulmonary Unit - A.O.U “Mater Domini”, Catanzaro;
3. Respiratory Medicine Unit - A.O.U. “Policlinico Giaccone”, Palermo;
4. Allergology and Clinical Immunology - University Hospital of Foggia, Foggia;
5. Department of Pneumology, A.O.R.N. “Dei Colli”, Naples;
6. Respiratory Medicine Unit - A.O.U. Policlinico di Bari “Giovanni XXIII”, Bari;
7. Respiratory Medicine Unit, A.O.U. “San Giovanni di Dio and Ruggi d’Aragona”, Salerno;
8. Allergology and Clinical Immunology - A.O.U. “San Giovanni di Dio and Ruggi d'Aragona”, Salerno;
9. Allergology, A.O.U. Policlinico di Bari “Giovanni XXIII”, Bari;
10. Institute of Respiratory Diseases - University Hospital of Foggia, Foggia;
11. Allergy and Pulmonary Medicine Unit - Center for Severe Asthma - ASP Palermo, Palermo.

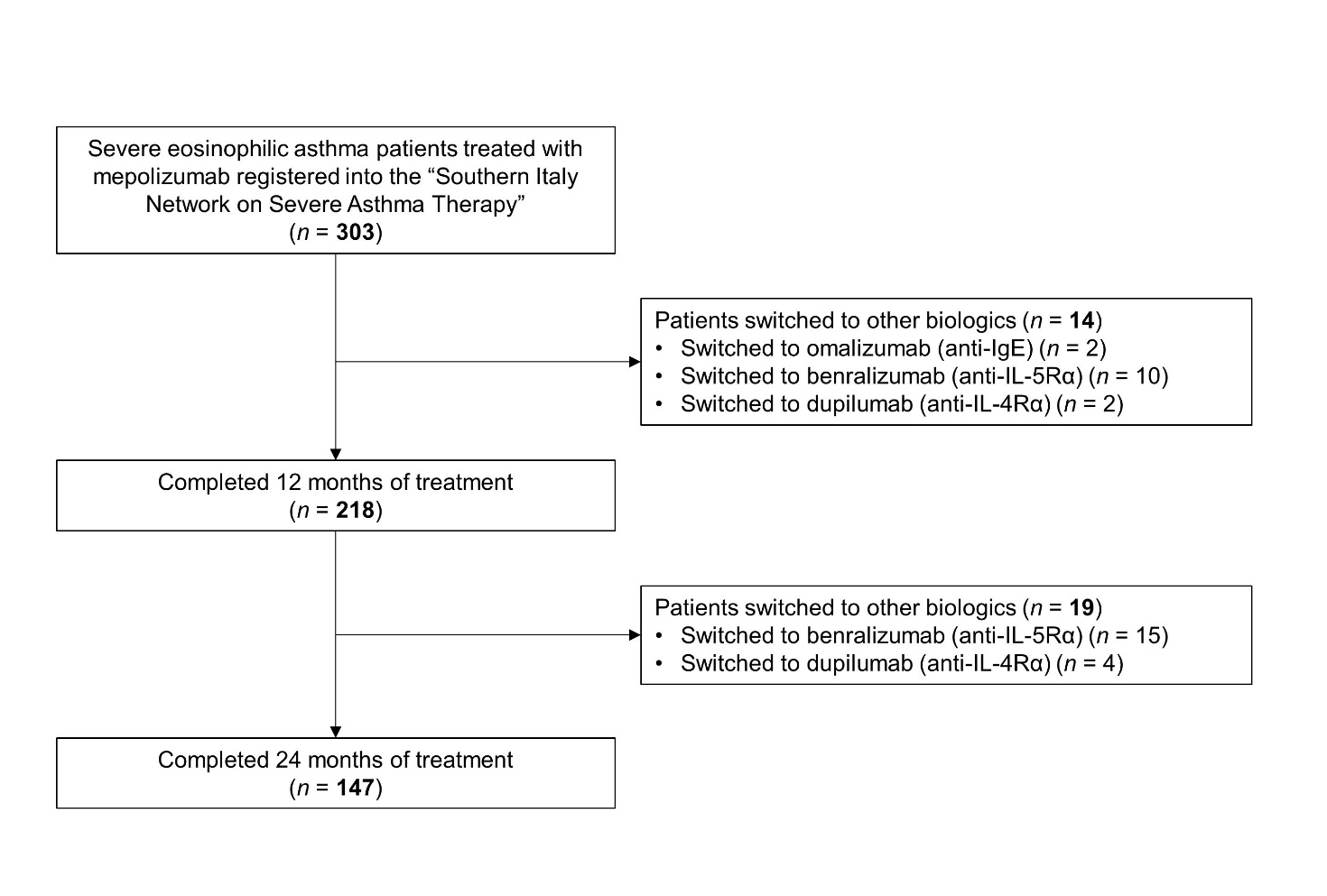
**Figure 1.** Study flow diagram.

. **TABLE E1.** Overall effects of mepolizumab therapy

|  | **n** | **Baseline** | **n** | **3 months** | ***P-value**** | **n** | **6 months** | ***P-value**** | **n** | **12 months** | ***P-value**** | **n** | **24 months** | ***P-value**** |
| --- | --- | --- | --- | --- | --- | --- | --- | --- | --- | --- | --- | --- | --- | --- |
| **Asthma outcomes** |  |  |  |  |  |  |  |  |  |  |  |  |  |  |
| Exacerbations / year, median (IQR) | 285 | 4 (2-6) | - | - | - | - | - | - | 218 | 1 (0-2) | **<0.0001** | 147 | 0 (0-1) | **<0.0001** |
| Patients who required ER / hospitalization, n (%) | 285 | 83 (29.1) | - | - | - | - | - | - | 218 | 20 (9.2) | **<0.0001** | 147 | 3 (2) | **<0.0001** |
| ACT, median (IQR) | 288 | 13 (10-17) | 219 | 21 (18-23) | **<0.0001** | 228 | 21 (18-23) | **<0.0001** | 215 | 21 (19-24) | **<0.0001** | 141 | 22 (20-24) | **<0.0001** |
| FEV_1_, %, median (IQR) | 290 | 71 (57-85) | 203 | 80.9 (64-98) | **<0.0001** | 213 | 83 (66.5-97) | **<0.0001** | 201 | 80 (68-97) | **<0.0001** | 128 | 80 (68-95) | **0.0001** |
| FEV_1_, L, median (IQR) | 289 | 1.8 (1.3-2.4) | 201 | 2.02 (1.6-2.6) | **<0.0001** | 210 | 2.09 (1.6-2.7) | **<0.0001** | 199 | 2.06 (1.6-2.8) | **<0.0001** | 123 | 2.0 (1.6-2.6) | **0.0051** |
| FVC, %, median (IQR) | 281 | 88 (73.5-100) | 192 | 94 (80-106) | **<0.0001** | 203 | 94 (80-106) | **<0.0001** | 190 | 94 (80-107) | **<0.0001** | 128 | 92 (80-103) | 0.0797 |
| FEV_1_/FVC, %, median (IQR) | 280 | 67 (58.5-76) | 190 | 72 (64-84) | **<0.0001** | 213 | 73 (63.5-84) | **<0.0001** | 201 | 73 (62-81) | **0.0003** | 128 | 72 (64-83) | **0.0016** |
| FEF_25-75_, %, median (IQR) | 228 | 37.5 (24-56.6) | 154 | 48 (29-68) | **<0.0001** | 149 | 46 (27.8-70) | **<0.0001** | 145 | 48 (29-69) | **<0.0001** | 128 | 50 (31-72) | **<0.0001** |
| **Pharmacologic therapies** |  |  |  |  |  |  |  |  |  |  |  |  |  |  |
| Patients on OCS, n, (%) | 294 | 204 (69.4) | 223 | 75 (33.6) | **<0.0001** | 223 | 58 (26) | **<0.0001** | 208 | 39 (18.8) | **<0.0001** | 139 | 23 (16.5) | **<0.0001** |
| OCS, mg/day, median (IQR) | 294 | 10 (2.5-25) | 223 | 2.3 (0-10) | **<0.0001** | 223 | 0 (0-5) | **<0.0001** | 208 | 0 (0-0.6) | **<0.0001** | 139 | 0 (0-0) | **<0.0001** |
| **Biomarkers** |  |  |  |  |  |  |  |  |  |  |  |  |  |  |
| Blood eosinophils, cells/μL median (IQR) | 282 | 490 (347-836) | 180 | 80 (40-125) | **<0.0001** | 183 | 79 (50-122) | **<0.0001** | 174 | 80 (50-130) | **<0.0001** | 140 | 80 (60-110) | **<0.0001** |
| Blood basophils, cells/μL median (IQR) | 122 | 50 (30-80) | 82 | 34 (20-50) | **0.0177** | 87 | 40 (20-60) | 0.1316 | 94 | 30 (19-60) | **0.0234** | 79 | 28 (20-57) | **0.0004** |
| FeNO, ppb, median (IQR) | 191 | 30 (14-55) | 98 | 20.5 (11-41) | 0.2911 | 125 | 26 (16-50) | 0.9876 | 115 | 25 (15-45) | 0.4260 | 79 | 25.5 (15-37.5) | 0.2416 |

Data are presented as mean (SD), n (%) or median (interquartile range). Bold highlights statistically significant *P-values*. *Abbreviations: ACT: Asthma Control Test; FEV1: forced expiratory volume in 1 s; FVC: forced vital capacity; FEF_25–75_%: forced expiratory flow 25–75%; OCS: oral corticosteroids; SNOT-22: Sino Nasal Outcome Test 22; OCS: Oral Corticosteroids; FeNO: Fraction of Exhaled Nitric Oxide*

*for comparisons vs baseline

**Table E2.** Rates of clinical and sustained remission after 12 and 24 months of treatment

|  | **n** | **12 months** | **n** | **24 months** | **n** | **Sustained from month 12 to 24** |
| --- | --- | --- | --- | --- | --- | --- |
| Zero exacerbations + zero OCS, n (%) | 206 | 98 (47.6) | 138 | 84 (60.9) | 138 | 45 (32.6) |
| **Remission criteria** |  |  |  |  |  |  |
| Zero exacerbations + zero OCS + ACT ≥20, n (%) | 206 | 89 (43.2) | 138 | 73 (52.9) | 138 | 40 (29) |
| Zero exacerbations + zero OCS + ACT ≥20 + FEV_1_ ≥80%, n (%) | 201 | 63 (31.3) | 128 | 50 (39.1) | 128 | 28 (21.9) |
| Zero exacerbations + zero OCS + ACT ≥20 + FEV_1_ +100mL from baseline, n (%) | 199 | 57 (28.6) | 123 | 42 (34.1) | 123 | 20 (16.3) |
| Zero exacerbations + zero OCS + ACT ≥20 + FEV_1_ decline ≤ 5% from baseline, n (%) | 201 | 72 (35.8) | 128 | 60 (46.9) | 128 | 31 (24.2) |
| Zero exacerbations + zero OCS + ACT ≥20 + FEV_1_ decline <100mL from the best value of the first 12 months. | - | - | 123 | 33 (26.8) | 123 | 18 (14.6) |

*Abbreviations: OCS: oral corticosteroids; ACT: Asthma Control Test; FEV_1_: forced expiratory volume in 1 s.*

**Table E3.** Remission criteria: Zero exacerbations + zero OCS + ACT ≥20.

Baseline characteristics comparing patients achieving remission vs non-remitters after 24 months of treatment

| **Zero exacerbations + zero OCS + ACT ≥20** | **n** | **Remission** | **n** | **No remission** | ***P-value*** |
| --- | --- | --- | --- | --- | --- |
| **General characteristics** |  |  |  |  |  |
| Age, years, mean (SD) | 73 | 57.1 (12) | 65 | 58.7 (10.4) | 0.4597 |
| Female, n (%) | 73 | 42 (57.5) | 65 | 48 (73.9) | 0.0506 |
| BMI, mean (SD) | 73 | 27 (5.7) | 65 | 27.9 (6.2) | 0.5125 |
| Obese (BMI ≥ 30), n (%) | 73 | 18 (24.7) | 65 | 19 (29.2) | 0.5689 |
| Duration of disease, years, median (IQR) | 73 | 18 (10-27) | 65 | 20.5 (10.7-32.5) | 0.2852 |
| Age at onset, years, median (IQR) | 73 | 37 (26-43) | 65 | 35.5 (24.5-50) | 0.5294 |
| Patients with positive Skin Prick Tests, n (%) | 73 | 48 (65.8) | 65 | 42 (64.6) | 0.9999 |
| Patients with positive Skin Prick Tests (perennial aeroallergens), n (%) | 73 | 26 (35.6) | 65 | 20 (30) | 0.5905 |
| Smoking status | 73 | 15 (20.6) | 65 | 26 (40) | **0.0155** |
| Smoking history, n (%) | 73 | 13 (17.8) | 65 | 20 (30.8) | 0.1090 |
| Current smoker, n (%) | 73 | 2 (2.7) | 65 | 6 (9.2) | 0.1479 |
| **Comorbidities** |  |  |  |  |  |
| Patients with anxiety/depression, n (%) | 73 | 3 (4.1) | 65 | 13 (20) | **0.0062** |
| Patients with GERD, n (%) | 73 | 16 (21.9) | 65 | 28 (43.1) | **0.0103** |
| Patients with bronchiectasis, n (%) | 73 | 12 (16.4) | 65 | 17 (26.5) | 0.2097 |
| Patients with atopic dermatitis, n (%) | 73 | 2 (2.7) | 65 | 3 (4.6) | 0.6662 |
| Patients with AERD, n (%) | 73 | 9 (12.3) | 65 | 4 (6.2) | 0.2544 |
| Patients with osteoporosis, n (%) | 73 | 7 (9.6) | 65 | 9 (13.9) | 0.5955 |
| Patients with CRSwNP, n (%) | 73 | 30 (47.6) | 65 | 29 (44.6) | 0.8593 |
| **Asthma outcomes** |  |  |  |  |  |
| Exacerbations / year, median (IQR) | 73 | 4 (2.3-8) | 65 | 5 (3.3-6) | 0.3364 |
| Patients who required ER / hospitalization, n (%) | 73 | 17 (23.3) | 65 | 22 (33.8) | 0.1888 |
| ACT, median (IQR) | 73 | 13.5 (10.8-18) | 65 | 12 (9.8-15) | 0.1343 |
| FEV_1_, %, median (IQR) | 68 | 71 (56-88) | 60 | 65 (48.5-83.5) | 0.1599 |
| FEV_1_, L, median (IQR) | 73 | 1.8 (1.3-2.4) | 65 | 1.6 (1.2-2.0) | 0.1235 |
| FVC, %, median (IQR) | 68 | 88 (71-100) | 60 | 83 (68.3-94.5) | 0.4895 |
| FEV_1_/FVC, %, median (IQR) | 68 | 73 (59-78.6) | 60 | 65.6 (57.4-72) | **0.0149** |
| FEF_25-75_, %, median (IQR) | 68 | 37.2 (25.8-58.3) | 60 | 26.5 (15.2-51.1) | **0.0219** |
| **Pharmacologic therapies** |  |  |  |  |  |
| High dose ICS-LABA, n (%) | 73 | 73 (100) | 65 | 65 (100) | 0.9999 |
| LAMA, n (%) | 73 | 39 (53.4) | 65 | 48 (73.9) | **0.0143** |
| Patients on OCS, n, (%) | 73 | 42 (57.5) | 65 | 47 (72.3) | 0.0775 |
| OCS, mg/die, median (IQR) | 73 | 7.8 (0-25) | 65 | 18 (5-25) | **0.0280** |
| Previous mAbs, n (%) | 73 | 10 (13.7) | 65 | 11 (16.9) | 0.6409 |
| **Biomarkers** |  |  |  |  |  |
| Blood eosinophils, cells/μL median (IQR) | 73 | 654 (380-1030) | 65 | 510 (400-896) | 0.6290 |
| Blood basophils, cells/μL median (IQR) | 47 | 60 (40-100) | 32 | 44 (30-60) | **0.0355** |
| IgE, UI/ml, median (IQR) | 56 | 271 (84-417) | 48 | 173 (32.8-408) | 0.2132 |
| FeNO, ppb, median (IQR) | 48 | 46 (23-112.8) | 31 | 31.4 (16.5-48) | 0.0572 |

Data are presented as mean (SD), n (%) or median (interquartile range). *Abbreviations: BMI: body mass index, GERD: gastroesophageal reflux disease; AERD: aspirin-exacerbated respiratory disease; CRwNP: Chronic Rhinosinusitis with Nasal Polyps; ACT: Asthma Control Test; FEV_1_: forced expiratory volume in 1 s; FVC: forced vital capacity; FEF_25–75_%: forced expiratory flow 25–75%; ICS-LABA: inhaled corticosteroids- long-acting β-agonists; LAMA: long acting muscarinic agonists; OCS: oral corticosteroids; mAb: monoclonal antibody; IgE, immunoglobulin-E; FeNO: Fraction of Exhaled Nitric Oxide.* Bold entries highlight statistically significant *P*-values.

**Table E4.** Remission criteria: Zero exacerbations + zero OCS + ACT ≥20.

Baseline characteristics comparing patients achieving sustained remission from month 12 to 24 vs non-remitters.

| **Zero exacerbations + zero OCS + ACT ≥20** | **n** | **Sustained remission** | **n** | **No remission** | ***P-value*** |
| --- | --- | --- | --- | --- | --- |
| **General characteristics** |  |  |  |  |  |
| Age, years, mean (SD) | 40 | 55.8 (9.7) | 98 | 58.7 (11.7) | 0.0735 |
| Female, n (%) | 40 | 23 (57.5) | 98 | 67 (68.4) | 0.2421 |
| BMI, mean (SD) | 40 | 27 (5.7) | 98 | 27.6 (6.0) | 0.4269 |
| Obese (BMI ≥ 30), n (%) | 40 | 8 (20) | 98 | 29 (29.6) | 0.2942 |
| Duration of disease, years, median (IQR) | 40 | 18 (11-28) | 98 | 18 (10-30) | 0.7433 |
| Age at onset, years, median (IQR) | 40 | 33 (25-43) | 98 | 38.5 (25.8-49.3) | 0.1760 |
| Patients with positive Skin Prick Tests, n (%) | 40 | 28 (70) | 98 | 62 (63.3) | 0.5555 |
| Patients with positive Skin Prick Tests (perennial aeroallergens), n (%) | 40 | 13 (32.5) | 98 | 33 (33.7) | 0.9999 |
| Smoking status | 40 | 8 (20) | 98 | 33 (33.7) | 0.1505 |
| Smoking history, n (%) | 40 | 8 (20) | 98 | 25 (25.5) | 0.6604 |
| Current smoker, n (%) | 40 | 0 (0) | 98 | 8 (8.2) | 0.1046 |
| **Comorbidities** |  |  |  |  |  |
| Patients with anxiety/depression, n (%) | 40 | 1 (2.5) | 98 | 15 (15.3) | **0.0392** |
| Patients with GERD, n (%) | 40 | 11 (27.5) | 98 | 33 (33.7) | 0.5494 |
| Patients with bronchiectasis, n (%) | 40 | 4 (10) | 98 | 25 (25.5) | 0.0636 |
| Patients with atopic dermatitis, n (%) | 40 | 0 (0) | 98 | 5 (5.1) | 0.3212 |
| Patients with AERD, n (%) | 40 | 5 (12.5) | 98 | 8 (8.2) | 0.5218 |
| Patients with osteoporosis, n (%) | 40 | 2 (5) | 98 | 14 (14.3) | 0.1513 |
| Patients with CRSwNP, n (%) | 40 | 20 (50) | 98 | 39 (39.8) | 0.3433 |
| **Asthma outcomes** |  |  |  |  |  |
| Exacerbations / year, median (IQR) | 40 | 4 (2-6.5) | 98 | 5 (3-6) | 0.2889 |
| Patients who required ER / hospitalization, n (%) | 40 | 11 (27.5) | 98 | 29 (29.6) | 0.8397 |
| ACT, median (IQR) | 40 | 13.5 (10-18) | 98 | 12.5 (9.8-15) | 0.5047 |
| FEV_1_, %, median (IQR) | 38 | 75 (52.8-88) | 90 | 65 (54-82) | 0.1278 |
| FEV_1_, L, median (IQR) | 40 | 1.9 (1.4-2.7) | 98 | 1.6 (1.2-2.0) | **0.0278** |
| FVC, %, median (IQR) | 38 | 89 (72.3-101) | 90 | 84 (68-97) | 0.4427 |
| FEV_1_/FVC, %, median (IQR) | 38 | 73.7 (64.6-81) | 90 | 66 (57.6-74.9) | **0.0019** |
| FEF_25-75_, %, median (IQR) | 38 | 41 (27-59) | 90 | 29 (16-51) | **0.0160** |
| **Pharmacologic therapies** |  |  |  |  |  |
| High dose ICS-LABA, n (%) | 40 | 40 (100) | 98 | 98 (100) | 0.9999 |
| LAMA, n (%) | 40 | 22 (55) | 98 | 65 (66.3) | 0.2455 |
| Patients on OCS, n, (%) | 40 | 23 (57.5) | 98 | 66 (67.4) | 0.3279 |
| OCS, mg/die, median (IQR) | 40 | 5 (0-12.5) | 98 | 12.5 (4-25) | **0.0047** |
| Previous mAbs, n (%) | 40 | 7 (17.5) | 98 | 14 (14.3) | 0.6115 |
| **Biomarkers** |  |  |  |  |  |
| Blood eosinophils, cells/μL median (IQR) | 40 | 685 (395-1059) | 98 | 520 (380-850) | 0.2785 |
| Blood basophils, cells/μL median (IQR) | 23 | 60 (48-100) | 52 | 50 (30-86) | 0.0728 |
| IgE, UI/ml, median (IQR) | 27 | 285 (111-422) | 77 | 167 (40-403) | 0.0895 |
| FeNO, ppb, median (IQR) | 27 | 61 (28.8-124.3) | 48 | 32 (13.9-55) | **0.0149** |

Data are presented as mean (SD), n (%) or median (interquartile range). *Abbreviations: BMI: body mass index, GERD: gastroesophageal reflux disease; AERD: aspirin-exacerbated respiratory disease; CRwNP: Chronic Rhinosinusitis with Nasal Polyps; ACT: Asthma Control Test; FEV_1_: forced expiratory volume in 1 s; FVC: forced vital capacity; FEF_25–75_%: forced expiratory flow 25–75%; ICS-LABA: inhaled corticosteroids- long-acting β-agonists; LAMA: long acting muscarinic agonists; OCS: oral corticosteroids; mAb: monoclonal antibody; IgE, immunoglobulin-E; FeNO: Fraction of Exhaled Nitric Oxide.* Bold entries highlight statistically significant *P*-values.

**Table E5.** Remission criteria: Zero exacerbations + zero OCS + ACT ≥20 + FEV_1_ ≥80%.

Baseline characteristics comparing patients achieving remission vs non-remitters after 24 months of treatment

| **Zero exacerbations + zero OCS + ACT ≥20 + FEV_1_ ≥80%** | **n** | **Remission** | **n** | **No remission** | ***P-value*** |
| --- | --- | --- | --- | --- | --- |
| **General characteristics** |  |  |  |  |  |
| Age, years, mean (SD) | 50 | 58.2 (11.5) | 78 | 57.9 (11.2) | 0.9102 |
| Female, n (%) | 50 | 30 (60) | 78 | 58 (74.3) | 0.1177 |
| BMI, mean (SD) | 50 | 26.8 (5.5) | 78 | 27.4 (5.4) | 0.5125 |
| Obese (BMI ≥ 30), n (%) | 50 | 14 (28) | 78 | 21 (26.9) | 0.9999 |
| Duration of disease, years, median (IQR) | 50 | 18.5 (11-28) | 78 | 18 (10.8-30) | 0.8839 |
| Age at onset, years, median (IQR) | 50 | 35.5 (26.3-42) | 78 | 39 (23-49) | 0.6421 |
| Patients with positive Skin Prick Tests, n (%) | 50 | 35 (70) | 78 | 52 (66.7) | 0.8462 |
| Patients with positive Skin Prick Tests (perennial aeroallergens), n (%) | 50 | 15 (30) | 78 | 31 (39.7) | 0.3454 |
| Smoking status | 50 | 10 (20) | 78 | 31 (39.7) | **0.0211** |
| Smoking history, n (%) | 50 | 8 (16) | 78 | 25 (32.1) | 0.0615 |
| Current smoker, n (%) | 50 | 2 (4) | 78 | 6 (7.7) | 0.4808 |
| **Comorbidities** |  |  |  |  |  |
| Patients with anxiety/depression, n (%) | 50 | 1 (2) | 78 | 18 (23.1) | **0.0007** |
| Patients with GERD, n (%) | 50 | 11 (22) | 78 | 29 (37.2) | 0.0810 |
| Patients with bronchiectasis, n (%) | 50 | 9 (18) | 78 | 18 (23.1) | 0.6575 |
| Patients with atopic dermatitis, n (%) | 50 | 1 (2) | 78 | 4 (5.1) | 0.6478 |
| Patients with AERD, n (%) | 50 | 8 (16) | 78 | 3 (3.9) | **0.0234** |
| Patients with osteoporosis, n (%) | 50 | 6 (12) | 78 | 9 (11.5) | 0.9999 |
| Patients with CRSwNP, n (%) | 50 | 22 (44) | 78 | 27 (34.6) | 0.3520 |
| **Asthma outcomes** |  |  |  |  |  |
| Exacerbations / year, median (IQR) | 50 | 4 (2-6) | 78 | 5 (3-8) | 0.0557 |
| Patients who required ER / hospitalization, n (%) | 50 | 13 (26) | 78 | 24 (30.8) | 0.6899 |
| ACT, median (IQR) | 50 | 14 (11-18) | 78 | 12 (9-15) | **0.0475** |
| FEV_1_, %, median (IQR) | 50 | 75.2 (61.5-90) | 78 | 65 (50.5-82) | **0.0066** |
| FEV_1_, L, median (IQR) | 50 | 2.0 (1.4-2.6) | 78 | 1.6 (1.3-2.1) | **0.0458** |
| FVC, %, median (IQR) | 50 | 92 (73.3-103.3) | 78 | 84 (68-95.8) | 0.0790 |
| FEV_1_/FVC, %, median (IQR) | 50 | 73.2 (59.7-78.4) | 78 | 67.5 (58-75.4) | 0.1151 |
| FEF_25-75_, %, median (IQR) | 50 | 45 (30.5-60) | 78 | 26.3 (16.9-46.8) | **0.0005** |
| **Pharmacologic therapies** |  |  |  |  |  |
| High dose ICS-LABA, n (%) | 50 | 50 (100) | 78 | 78 (100) | 0.9999 |
| LAMA, n (%) | 50 | 25 (50) | 78 | 62 (79.5) | **0.0008** |
| Patients on OCS, n, (%) | 50 | 27 (54) | 78 | 60 (76.9) | **0.0111** |
| OCS, mg/die, median (IQR) | 50 | 8.8 (0-25) | 78 | 12.5 (2.9-25) | 0.3486 |
| Previous mAbs, n (%) | 50 | 9 (18) | 78 | 12 (15.4) | 0.8076 |
| **Biomarkers** |  |  |  |  |  |
| Blood eosinophils, cells/μL median (IQR) | 50 | 630 (387-1090) | 78 | 510 (387-858) | 0.4258 |
| Blood basophils, cells/μL median (IQR) | 24 | 61 (42.8-97.5) | 55 | 50 (30-86) | 0.0997 |
| IgE, UI/ml, median (IQR) | 33 | 275 (93-435) | 71 | 167 (42-423) | 0.2690 |
| FeNO, ppb, median (IQR) | 29 | 52 (21.5-106.4) | 50 | 31.2 (19.2-51.2) | 0.0833 |

Data are presented as mean (SD), n (%) or median (interquartile range). *Abbreviations: BMI: body mass index, GERD: gastroesophageal reflux disease; AERD: aspirin-exacerbated respiratory disease; CRwNP: Chronic Rhinosinusitis with Nasal Polyps; ACT: Asthma Control Test; FEV_1_: forced expiratory volume in 1 s; FVC: forced vital capacity; FEF_25–75_%: forced expiratory flow 25–75%; ICS-LABA: inhaled corticosteroids- long-acting β-agonists; LAMA: long acting muscarinic agonists; OCS: oral corticosteroids; mAb: monoclonal antibody; IgE, immunoglobulin-E; FeNO: Fraction of Exhaled Nitric Oxide.* Bold entries highlight statistically significant *P*-values.

**Table E6.** Remission criteria: Zero exacerbations + zero OCS + ACT ≥20 + FEV_1_ ≥80%.

Baseline characteristics comparing patients achieving sustained remission from month 12 to 24 vs non-remitters.

| **Zero exacerbations + zero OCS + ACT ≥20 + FEV_1_ ≥80%** | **n** | **Sustained remission** | **n** | **No remission** | ***P-value*** |
| --- | --- | --- | --- | --- | --- |
| **General characteristics** |  |  |  |  |  |
| Age, years, mean (SD) | 28 | 56.1 (10.5) | 100 | 58.5 (11.4) | 0.1705 |
| Female, n (%) | 28 | 16 (57.1) | 100 | 72 (72) | 0.1671 |
| BMI, mean (SD) | 28 | 26.5 (6.0) | 100 | 27.4 (5.3) | 0.2489 |
| Obese (BMI ≥ 30), n (%) | 28 | 6 (21.4) | 100 | 29 (29) | 0.4820 |
| Duration of disease, years, median (IQR) | 28 | 18 (10-28) | 100 | 19.5 (11-30) | 0.4570 |
| Age at onset, years, median (IQR) | 28 | 34 (26-43) | 100 | 39 (23.5-49) | 0.5369 |
| Patients with positive Skin Prick Tests, n (%) | 28 | 17 (60.7) | 100 | 70 (70) | 0.3670 |
| Patients with positive Skin Prick Tests (perennial aeroallergens), n (%) | 28 | 7 (25) | 100 | 39 (39) | 0.1900 |
| Smoking status | 28 | 7 (25) | 100 | 34 (34) | 0.4928 |
| Smoking history, n (%) | 28 | 7 (25) | 100 | 26 (26) | 0.9999 |
| Current smoker, n (%) | 28 | 0 (0) | 100 | 8 (8) | 0.1988 |
| **Comorbidities** |  |  |  |  |  |
| Patients with anxiety/depression, n (%) | 28 | 1 (3.5) | 100 | 18 (18) | 0.0720 |
| Patients with GERD, n (%) | 28 | 6 (21.4) | 100 | 34 (34) | 0.2530 |
| Patients with bronchiectasis, n (%) | 28 | 1 (3.6) | 100 | 26 (26) | **0.0083** |
| Patients with atopic dermatitis, n (%) | 28 | 0 (0) | 100 | 5 (5) | 0.5850 |
| Patients with AERD, n (%) | 28 | 3 (10.7) | 100 | 8 (8) | 0.3770 |
| Patients with osteoporosis, n (%) | 28 | 2 (7.1) | 100 | 13 (13) | 0.5202 |
| Patients with CRSwNP, n (%) | 28 | 14 (50) | 100 | 35 (35) | 0.1877 |
| **Asthma outcomes** |  |  |  |  |  |
| Exacerbations / year, median (IQR) | 28 | 4 (2-6) | 100 | 5 (3-8) | 0.1318 |
| Patients who required ER / hospitalization, n (%) | 28 | 6 (21.4) | 100 | 31 (31) | 0.3580 |
| ACT, median (IQR) | 28 | 15 (11-18) | 100 | 12 (10-15) | 0.0785 |
| FEV_1_, %, median (IQR) | 28 | 83 (73.5-91) | 100 | 65 (51-80) | **0.0007** |
| FEV_1_, L, median (IQR) | 28 | 2.3 (1.6-2.8) | 100 | 1.6 (1.2-2.1) | **0.0014** |
| FVC, %, median (IQR) | 28 | 92 (79.6-100) | 100 | 84 (68-100) | **0.0494** |
| FEV_1_/FVC, %, median (IQR) | 28 | 74.3 (65.5-79) | 100 | 67 (58-75.8) | **0.0160** |
| FEF_25-75_, %, median (IQR) | 28 | 56 (40-75) | 100 | 29 (18-49) | **<0.0001** |
| **Pharmacologic therapies** |  |  |  |  |  |
| High dose ICS-LABA, n (%) | 28 | 28 (100) | 100 | 100 (100) | 0.9999 |
| LAMA, n (%) | 28 | 12 (42.9) | 100 | 75 (75) | **0.0024** |
| Patients on OCS, n, (%) | 28 | 13 (46.4) | 100 | 74 (74) | **0.0106** |
| OCS, mg/die, median (IQR) | 28 | 6.4 (0-25) | 100 | 12.5 (2.5-25) | 0.1208 |
| Previous mAbs, n (%) | 28 | 5 (17.9) | 100 | 16 (16) | 0.7787 |
| **Biomarkers** |  |  |  |  |  |
| Blood eosinophils, cells/μL median (IQR) | 28 | 545 (372-1058) | 100 | 545 (390-880) | 0.8361 |
| Blood basophils, cells/μL median (IQR) | 15 | 60 (41-90) | 64 | 50 (30-90) | 0.5421 |
| IgE, UI/ml, median (IQR) | 18 | 276 (98-424) | 86 | 173 (52-418) | 0.3655 |
| FeNO, ppb, median (IQR) | 16 | 66 (30-124) | 63 | 31.7 (15.8-56) | **0.0113** |

Data are presented as mean (SD), n (%) or median (interquartile range). *Abbreviations: BMI: body mass index, GERD: gastroesophageal reflux disease; AERD: aspirin-exacerbated respiratory disease; CRwNP: Chronic Rhinosinusitis with Nasal Polyps; ACT: Asthma Control Test; FEV_1_: forced expiratory volume in 1 s; FVC: forced vital capacity; FEF_25–75_%: forced expiratory flow 25–75%; ICS-LABA: inhaled corticosteroids- long-acting β-agonists; LAMA: long acting muscarinic agonists; OCS: oral corticosteroids; mAb: monoclonal antibody; IgE, immunoglobulin-E; FeNO: Fraction of Exhaled Nitric Oxide.* Bold entries highlight statistically significant *P*-values.

**Table E7.** Remission criteria: Zero exacerbations + zero OCS + ACT ≥20 + FEV_1_ +100mL from baseline.

Baseline characteristics comparing patients achieving remission vs non-remitters after 24 months of treatment.

| **Zero exacerbations + zero OCS + ACT ≥20 + FEV_1_ +100mL** | **n** | **Remission** | **n** | **No remission** | ***P-value*** |
| --- | --- | --- | --- | --- | --- |
| **General characteristics** |  |  |  |  |  |
| Age, years, mean (SD) | 42 | 57.2 (13) | 81 | 58.1 (10.3) | 0.9882 |
| Female, n (%) | 42 | 23 (54.8) | 81 | 60 (74.1) | **0.0420** |
| BMI, mean (SD) | 42 | 27.2 (5) | 81 | 27.4 (5.8) | 0.8485 |
| Obese (BMI ≥ 30), n (%) | 42 | 13 (31) | 81 | 20 (24.7) | 0.5217 |
| Duration of disease, years, median (IQR) | 42 | 18 (10-27.5) | 81 | 17.5 (10-30) | 0.6933 |
| Age at onset, years, median (IQR) | 42 | 38 (25-44) | 81 | 40 (25.5-49) | 0.5062 |
| Patients with positive Skin Prick Tests, n (%) | 42 | 27 (64.3) | 81 | 55 (67.9) | 0.6918 |
| Patients with positive Skin Prick Tests (perennial aeroallergens), n (%) | 42 | 15 (35.7) | 81 | 26 (32.1) | 0.6918 |
| Smoking status | 42 | 6 (14.3) | 81 | 32 (39.5) | **0.0041** |
| Smoking history, n (%) | 42 | 6 (14.3) | 81 | 25 (30.9) | 0.0508 |
| Current smoker, n (%) | 42 | 0 (0) | 81 | 7 (8.6) | 0.0941 |
| **Comorbidities** |  |  |  |  |  |
| Patients with anxiety/depression, n (%) | 42 | 2 (4.8) | 81 | 16 (19.8) | **0.0308** |
| Patients with GERD, n (%) | 42 | 7 (16.7) | 81 | 30 (37) | **0.0229** |
| Patients with bronchiectasis, n (%) | 42 | 4 (9.5) | 81 | 20 (24.7) | 0.0553 |
| Patients with atopic dermatitis, n (%) | 42 | 1 (2.4) | 81 | 3 (3.7) | 0.9999 |
| Patients with AERD, n (%) | 42 | 2 (4.8) | 81 | 7 (8.6) | 0.7196 |
| Patients with osteoporosis, n (%) | 42 | 3 (7.1) | 81 | 9 (11.1) | 0.7497 |
| Patients with CRSwNP, n (%) | 42 | 18 (42.9) | 81 | 35 (43.2) | 0.9999 |
| **Asthma outcomes** |  |  |  |  |  |
| Exacerbations / year, median (IQR) | 42 | 4 (2-8) | 81 | 5 (3-7) | 0.1474 |
| Patients who required ER / hospitalization, n (%) | 42 | 11 (26.2) | 81 | 23 (28.4) | 0.8350 |
| ACT, median (IQR) | 42 | 13 (8.5-16) | 81 | 12 (10-15) | 0.8663 |
| FEV_1_, %, median (IQR) | 42 | 63 (51.3-83.2) | 81 | 69 (59-87) | 0.2714 |
| FEV_1_, L, median (IQR) | 42 | 1.6 (1.2-2.2) | 81 | 1.7 (1.4-2.3) | 0.5091 |
| FVC, %, median (IQR) | 42 | 79 (63-95) | 81 | 89 (74.6-101) | 0.0577 |
| FEV_1_/FVC, %, median (IQR) | 42 | 73.8 (59.4-80.5) | 81 | 69 (58-76.8) | 0.1230 |
| FEF_25-75_, %, median (IQR) | 42 | 33.5 (24.8-50) | 81 | 31.9 (18-56) | 0.5400 |
| **Pharmacologic therapies** |  |  |  |  |  |
| High dose ICS-LABA, n (%) | 42 | 42 (100) | 81 | 81 (100) | 0.9999 |
| LAMA, n (%) | 42 | 26 (61.9) | 81 | 56 (69.1) | 0.4279 |
| Patients on OCS, n, (%) | 42 | 26 (61.9) | 81 | 56 (69.1) | 0.4279 |
| OCS, mg/die, median (IQR) | 42 | 5 (2.5-12.5) | 81 | 12.5 (2.9-25) | 0.1246 |
| Previous mAbs, n (%) | 42 | 7 (16.7) | 81 | 14 (17.3) | 0.9999 |
| **Biomarkers** |  |  |  |  |  |
| Blood eosinophils, cells/μL median (IQR) | 42 | 550 (360-848) | 81 | 500 (382-807) | 0.7856 |
| Blood basophils, cells/μL median (IQR) | 23 | 53 (30-95) | 52 | 50 (32.5-80) | 0.3091 |
| IgE, UI/ml, median (IQR) | 27 | 257 (105-429) | 77 | 181 (42-431) | 0.6349 |
| FeNO, ppb, median (IQR) | 27 | 47.5 (28-114) | 48 | 30 (15-57) | 0.0595 |

Data are presented as mean (SD), n (%) or median (interquartile range). *Abbreviations: BMI: body mass index, GERD: gastroesophageal reflux disease; AERD: aspirin-exacerbated respiratory disease; CRwNP: Chronic Rhinosinusitis with Nasal Polyps; ACT: Asthma Control Test; FEV_1_: forced expiratory volume in 1 s; FVC: forced vital capacity; FEF_25–75_%: forced expiratory flow 25–75%; ICS-LABA: inhaled corticosteroids- long-acting β-agonists; LAMA: long acting muscarinic agonists; OCS: oral corticosteroids; mAb: monoclonal antibody; IgE, immunoglobulin-E; FeNO: Fraction of Exhaled Nitric Oxide.* Bold entries highlight statistically significant *P*-values.

| **Zero exacerbations + zero OCS + ACT ≥20 + FEV_1_ +100mL** | **n** | **Sustained remission** | **n** | **No remission** | ***P-value*** |
| --- | --- | --- | --- | --- | --- |
| **General characteristics** |  |  |  |  |  |
| Age, years, mean (SD) | 20 | 57.8 (11.4) | 103 | 57.8 (11.3) | 0.9651 |
| Female, n (%) | 20 | 10 (50) | 103 | 73 (70.9) | 0.1152 |
| BMI, mean (SD) | 20 | 26.1 (4.1) | 103 | 27.6 (5.7) | 0.4288 |
| Obese (BMI ≥ 30), n (%) | 20 | 3 (15) | 103 | 30 (29.1) | 0.2723 |
| Duration of disease, years, median (IQR) | 20 | 20 (14-25) | 103 | 17 (10-30) | 0.6216 |
| Age at onset, years, median (IQR) | 20 | 38 (25-43) | 103 | 39 (26-49) | 0.7062 |
| Patients with positive Skin Prick Tests, n (%) | 20 | 11 (55) | 103 | 71 (68.9) | 0.2998 |
| Patients with positive Skin Prick Tests (perennial aeroallergens), n (%) | 20 | 6 (30) | 103 | 35 (34) | 0.8011 |
| Smoking status | 20 | 2 (10) | 103 | 36 (35) | **0.0333** |
| Smoking history, n (%) | 20 | 2 (10) | 103 | 29 (28.2) | 0.0995 |
| Current smoker, n (%) | 20 | 0 (0) | 103 | 7 (6.8) | 0.5974 |
| **Comorbidities** |  |  |  |  |  |
| Patients with anxiety/depression, n (%) | 20 | 0 (0) | 103 | 18 (17.5) | **0.0421** |
| Patients with GERD, n (%) | 20 | 2 (10) | 103 | 35 (34) | **0.0347** |
| Patients with bronchiectasis, n (%) | 20 | 1 (5) | 103 | 23 (95.8) | 0.1195 |
| Patients with atopic dermatitis, n (%) | 20 | 0 (0) | 103 | 4 (3.9) | 0.9999 |
| Patients with AERD, n (%) | 20 | 0 (0) | 103 | 9 (8.7) | 0.3526 |
| Patients with osteoporosis, n (%) | 20 | 0 (0) | 103 | 12 (11.7) | 0.2121 |
| Patients with CRSwNP, n (%) | 20 | 8 (40) | 103 | 45 (43.7) | 0.8098 |
| **Asthma outcomes** |  |  |  |  |  |
| Exacerbations / year, median (IQR) | 20 | 5 (3-8) | 103 | 5 (3-6) | 0.9748 |
| Patients who required ER / hospitalization, n (%) | 20 | 5 (25) | 103 | 29 (28.2) | 0.9999 |
| ACT, median (IQR) | 20 | 12 (7.8-16) | 103 | 13 (10-15) | 0.2522 |
| FEV_1_, %, median (IQR) | 20 | 63 (50-82) | 103 | 69 (58-86) | 0.3574 |
| FEV_1_, L, median (IQR) | 20 | 1.5 (1.2-2.0) | 103 | 1.7 (1.3-2.3) | 0.3108 |
| FVC, %, median (IQR) | 20 | 75 (65-89) | 103 | 89 (72.5-101) | **0.0379** |
| FEV_1_/FVC, %, median (IQR) | 20 | 76.2 (68.8-81.5) | 103 | 69 (58-76.8) | **0.0157** |
| FEF_25-75_, %, median (IQR) | 20 | 33 (19.5-47.6) | 103 | 32 (20-55.8) | 0.8183 |
| **Pharmacologic therapies** |  |  |  |  |  |
| High dose ICS-LABA, n (%) | 20 | 20 (100) | 103 | 103 (100) | 0.9999 |
| LAMA, n (%) | 20 | 10 (50) | 103 | 72 (69.9) | 0.1186 |
| Patients on OCS, n, (%) | 20 | 12 (60) | 103 | 70 (68) | 0.6050 |
| OCS, mg/die, median (IQR) | 20 | 5 (2-18.8) | 103 | 12.5 (2.5-25) | 0.6327 |
| Previous mAbs, n (%) | 20 | 5 (25) | 103 | 16 (15.5) | 0.3330 |
| **Biomarkers** |  |  |  |  |  |
| Blood eosinophils, cells/μL median (IQR) | 20 | 670 (390-1145) | 103 | 500 (367-802) | 0.2928 |
| Blood basophils, cells/μL median (IQR) | 13 | 53 (26.8-111) | 62 | 50 (75-83) | 0.7285 |
| IgE, UI/ml, median (IQR) | 17 | 257 (105-457) | 87 | 181 (51-429) | 0.6161 |
| FeNO, ppb, median (IQR) | 16 | 69 (34.5-135) | 59 | 30 (16-58) | **0.0179** |

**Table E8.** Remission criteria: Zero exacerbations + zero OCS + ACT ≥20 + FEV1 +100mL from baseline.

Baseline characteristics comparing patients achieving sustained remission from month 12 to 24 vs non-remitters.

Data are presented as mean (SD), n (%) or median (interquartile range). *Abbreviations: BMI: body mass index, GERD: gastroesophageal reflux disease; AERD: aspirin-exacerbated respiratory disease; CRwNP: Chronic Rhinosinusitis with Nasal Polyps; ACT: Asthma Control Test; FEV_1_: forced expiratory volume in 1 s; FVC: forced vital capacity; FEF_25–75_%: forced expiratory flow 25–75%; ICS-LABA: inhaled corticosteroids- long-acting β-agonists; LAMA: long acting muscarinic agonists; OCS: oral corticosteroids; mAb: monoclonal antibody; IgE, immunoglobulin-E; FeNO: Fraction of Exhaled Nitric Oxide.* Bold entries highlight statistically significant *P*-values.

**Table E9.** Remission criteria: Zero exacerbations + zero OCS + ACT ≥20 + FEV_1_ decline ≤5% from baseline.

Baseline characteristics comparing patients achieving remission vs non-remitters after 24 months of treatment.

| **Zero exacerbations + zero OCS + ACT ≥20 +** **FEV_1_ decline ≤5% from baseline** | **n** | **Remission** | **n** | **No remission** | ***P-value*** |
| --- | --- | --- | --- | --- | --- |
| **General characteristics** |  |  |  |  |  |
| Age, years, mean (SD) | 60 | 57.5 (12.4) | 68 | 58.7 (10.6) | 0.6974 |
| Female, n (%) | 60 | 34 (56.7) | 68 | 54 (79.4) | **0.0074** |
| BMI, mean (SD) | 60 | 27.1 (5.7) | 68 | 27.3 (5.3) | 0.7401 |
| Obese (BMI ≥ 30), n (%) | 60 | 14 (23.3) | 68 | 21 (30.9) | 0.4274 |
| Duration of disease, years, median (IQR) | 60 | 18 (11-27) | 68 | 18 (10-30) | 0.6876 |
| Age at onset, years, median (IQR) | 60 | 37 (25-43) | 68 | 40 (25-50) | 0.3349 |
| Patients with positive Skin Prick Tests, n (%) | 60 | 39 (65) | 68 | 48 (70.6) | 0.5706 |
| Patients with positive Skin Prick Tests (perennial aeroallergens), n (%) | 60 | 18 (30) | 68 | 28 (41.2) | 0.2020 |
| Smoking status | 60 | 13 (21.7) | 68 | 28 (41.2) | **0.0229** |
| Smoking history, n (%) | 60 | 12 (20) | 68 | 21 (30.9) | 0.2242 |
| Current smoker, n (%) | 60 | 1 (1.7) | 68 | 7 (10.3) | 0.0660 |
| **Comorbidities** |  |  |  |  |  |
| Patients with anxiety/depression, n (%) | 60 | 0 (0) | 68 | 18 (17.5) | **0.0421** |
| Patients with GERD, n (%) | 60 | 13 (21.7) | 68 | 27 (39.7) | **0.0357** |
| Patients with bronchiectasis, n (%) | 60 | 6 (10) | 68 | 21 (30.9) | **0.0046** |
| Patients with atopic dermatitis, n (%) | 60 | 1 (1.7) | 68 | 4 (5.9) | 0.3701 |
| Patients with AERD, n (%) | 60 | 5 (8.3) | 68 | 6 (8.8) | 0.9999 |
| Patients with osteoporosis, n (%) | 60 | 5 (8.3) | 68 | 10 (14.7) | 0.2872 |
| Patients with CRSwNP, n (%) | 60 | 26 (43.3) | 68 | 23 (33.8) | 0.2811 |
| **Asthma outcomes** |  |  |  |  |  |
| Exacerbations / year, median (IQR) | 60 | 4 (2-8.3) | 68 | 5 (3.8-6) | 0.3037 |
| Patients who required ER / hospitalization, n (%) | 60 | 15 (25) | 68 | 22 (32.4) | 0.4358 |
| ACT, median (IQR) | 60 | 14 (10.3-16) | 68 | 13 (10-15) | 0.4340 |
| FEV_1_, %, median (IQR) | 60 | 69 (54-87) | 68 | 68.5 (57-85) | 0.8994 |
| FEV_1_, L, median (IQR) | 60 | 1.8 (1.3-2.4) | 68 | 1.6 (1.3-2.1) | 0.4295 |
| FVC, %, median (IQR) | 60 | 84 (65.1-95) | 68 | 86.5 (73-104) | 0.2352 |
| FEV_1_/FVC, %, median (IQR) | 60 | 74 (59.8-80) | 68 | 67 (58-74.6) | **0.0160** |
| FEF_25-75_, %, median (IQR) | 60 | 39.7 (25.8-56.2) | 68 | 28.5 (18-51.9) | 0.1344 |
| **Pharmacologic therapies** |  |  |  |  |  |
| High dose ICS-LABA, n (%) | 60 | 60 (100) | 68 | 68 (100) | 0.9999 |
| LAMA, n (%) | 60 | 33 (55) | 68 | 54 (79.4) | **0.0043** |
| Patients on OCS, n, (%) | 60 | 38 (63.3) | 68 | 59 (86.8) | **0.0034** |
| OCS, mg/die, median (IQR) | 60 | 7.8 (2.5-25) | 68 | 12.5 (0-25) | 0.3117 |
| Previous mAbs, n (%) | 60 | 10 (16.7) | 68 | 11 (16.2) | 0.9999 |
| **Biomarkers** |  |  |  |  |  |
| Blood eosinophils, cells/μL median (IQR) | 60 | 570 (362-960) | 68 | 500 (390-890) | 0.7859 |
| Blood basophils, cells/μL median (IQR) | 31 | 60 (41-91) | 48 | 44 (30-72) | **0.0496** |
| IgE, UI/ml, median (IQR) | 38 | 300 (108-462) | 66 | 151 (36.3-425) | 0.0908 |
| FeNO, ppb, median (IQR) | 32 | 46 (20-114) | 47 | 30.5 (16.3-48.5) | 0.0673 |

Data are presented as mean (SD), n (%) or median (interquartile range). *Abbreviations: BMI: body mass index, GERD: gastroesophageal reflux disease; AERD: aspirin-exacerbated respiratory disease; CRwNP: Chronic Rhinosinusitis with Nasal Polyps; ACT: Asthma Control Test; FEV_1_: forced expiratory volume in 1 s; FVC: forced vital capacity; FEF_25–75_%: forced expiratory flow 25–75%; ICS-LABA: inhaled corticosteroids- long-acting β-agonists; LAMA: long acting muscarinic agonists; OCS: oral corticosteroids; mAb: monoclonal antibody; IgE, immunoglobulin-E; FeNO: Fraction of Exhaled Nitric Oxide.* Bold entries highlight statistically significant *P*-values.

**Table E10.** Remission criteria: Zero exacerbations + zero OCS + ACT ≥20 + FEV_1_ decline ≤5% from baseline.

Baseline characteristics comparing patients achieving sustained remission from month 12 to 24 vs non-remitters.

| **Zero exacerbations + zero OCS + ACT ≥20 + FEV_1_ decline ≤5% from baseline** | **n** | **Sustained remission** | **n** | **No remission** | ***P-value*** |
| --- | --- | --- | --- | --- | --- |
| **General characteristics** |  |  |  |  |  |
| Age, years, mean (SD) | 31 | 55.9 (10.6) | 97 | 58.6 (11.6) | 0.1374 |
| Female, n (%) | 31 | 20 (64.5) | 97 | 68 (70.1) | 0.6569 |
| BMI, mean (SD) | 31 | 27.4 (6.5) | 97 | 27.1 (5.1) | 0.8063 |
| Obese (BMI ≥ 30), n (%) | 31 | 7 (22.6) | 97 | 28 (28.9) | 0.6444 |
| Duration of disease, years, median (IQR) | 31 | 18 (11.5-25) | 97 | 17.5 (10-30) | 0.7591 |
| Age at onset, years, median (IQR) | 31 | 35.5 (26.5-44.3) | 97 | 40 (25-49.5) | 0.3383 |
| Patients with positive Skin Prick Tests, n (%) | 31 | 20 (64.5) | 97 | 67 (69.1) | 0.6623 |
| Patients with positive Skin Prick Tests (perennial aeroallergens), n (%) | 31 | 8 (25) | 97 | 38 (39.2) | 0.2017 |
| Smoking status | 31 | 6 (19.4) | 97 | 35 (36.1) | 0.1207 |
| Smoking history, n (%) | 31 | 6 (19.4) | 97 | 27 (27.8) | 0.4800 |
| Current smoker, n (%) | 31 | 0 (0) | 97 | 8 (8.3) | 0.1974 |
| **Comorbidities** |  |  |  |  |  |
| Patients with anxiety/depression, n (%) | 31 | 0 (0) | 97 | 18 (17.5) | **0.0421** |
| Patients with GERD, n (%) | 31 | 7 (22.6) | 97 | 33 (34) | 0.2718 |
| Patients with bronchiectasis, n (%) | 31 | 0 (0) | 97 | 27 (27.8) | **0.0003** |
| Patients with atopic dermatitis, n (%) | 31 | 0 (0) | 97 | 5 (5.2) | 0.3349 |
| Patients with AERD, n (%) | 31 | 2 (6.5) | 97 | 9 (9.4) | 0.9999 |
| Patients with osteoporosis, n (%) | 31 | 1 (3.2) | 97 | 14 (14.4) | 0.1155 |
| Patients with CRSwNP, n (%) | 31 | 15 (48.4) | 97 | 34 (35.1) | 0.2070 |
| **Asthma outcomes** |  |  |  |  |  |
| Exacerbations / year, median (IQR) | 31 | 3.5 (2-7.5) | 97 | 5 (3-7) | 0.2191 |
| Patients who required ER / hospitalization, n (%) | 31 | 7 (22.6) | 97 | 30 (30.9) | 0.4958 |
| ACT, median (IQR) | 31 | 13 (8-16.5) | 97 | 13 (10.5-15) | 0.9254 |
| FEV_1_, %, median (IQR) | 31 | 78.5 (52.8-88.8) | 97 | 66 (57-85) | 0.2729 |
| FEV_1_, L, median (IQR) | 31 | 1.94 (1.4-2.6) | 97 | 1.6 (1.3-2.1) | 0.1488 |
| FVC, %, median (IQR) | 31 | 89 (72.3-96) | 97 | 85 (68-101) | 0.9095 |
| FEV_1_/FVC, %, median (IQR) | 31 | 77 (69.1-82.8) | 97 | 67 (58-75) | **0.0005** |
| FEF_25-75_, %, median (IQR) | 31 | 41.5 (28.4-73.5) | 97 | 29.5 (18-50) | **0.0274** |
| **Pharmacologic therapies** |  |  |  |  |  |
| High dose ICS-LABA, n (%) | 31 | 31 (100) | 97 | 97 (100) | 0.9999 |
| LAMA, n (%) | 31 | 15 (48.4) | 97 | 72 (74.2) | **0.0139** |
| Patients on OCS, n, (%) | 31 | 20 (64.5) | 97 | 60 (61.8) | 0.8344 |
| OCS, mg/die, median (IQR) | 31 | 5 (2.5-12.5) | 97 | 12.5 (2.5-25) | **0.0226** |
| Previous mAbs, n (%) | 31 | 6 (19.4) | 97 | 15 (15.5) | 0.5878 |
| **Biomarkers** |  |  |  |  |  |
| Blood eosinophils, cells/μL median (IQR) | 31 | 680 (398-1058) | 97 | 500 (358-802) | 0.1493 |
| Blood basophils, cells/μL median (IQR) | 17 | 60 (50-106) | 62 | 46.5 (30-78.8) | **0.0443** |
| IgE, UI/ml, median (IQR) | 19 | 335 (160-462) | 85 | 159 (47.3-427) | 0.0733 |
| FeNO, ppb, median (IQR) | 19 | 69 (30-124) | 60 | 31 (15-50) | **0.0029** |

Data are presented as mean (SD), n (%) or median (interquartile range). *Abbreviations: BMI: body mass index, GERD: gastroesophageal reflux disease; AERD: aspirin-exacerbated respiratory disease; CRwNP: Chronic Rhinosinusitis with Nasal Polyps; ACT: Asthma Control Test; FEV_1_: forced expiratory volume in 1 s; FVC: forced vital capacity; FEF_25–75_%: forced expiratory flow 25–75%; ICS-LABA: inhaled corticosteroids- long-acting β-agonists; LAMA: long acting muscarinic agonists; OCS: oral corticosteroids; mAb: monoclonal antibody; IgE, immunoglobulin-E; FeNO: Fraction of Exhaled Nitric Oxide.* Bold entries highlight statistically significant *P*-values.

**Table E11.** Remission criteria: Zero exacerbations + zero OCS + ACT ≥20 + FEV_1_ decline <100mL from the best value of the first 12 months. Baseline characteristics comparing patients achieving remission vs non-remitters after 24 months of treatment.

| **Zero exacerbations + zero OCS + ACT ≥20 +** **FEV_1_ decline <100mL from the best value of the first 12 months** | **n** | **Remission** | **n** | **No remission** | ***P-value*** |
| --- | --- | --- | --- | --- | --- |
| **General characteristics** |  |  |  |  |  |
| Age, years, mean (SD) | 33 | 58 (13.3) | 90 | 57.7 (10.5) | 0.6368 |
| Female, n (%) | 33 | 19 (57.6) | 90 | 64 (71.1) | 0.1933 |
| BMI, mean (SD) | 33 | 27.4 (5.6) | 90 | 27.3 (5.5) | 0.7621 |
| Obese (BMI ≥ 30), n (%) | 33 | 7 (21.2) | 90 | 26 (28.9) | 0.4936 |
| Duration of disease, years, median (IQR) | 33 | 18 (10-25) | 90 | 18 (10-30) | 0.4141 |
| Age at onset, years, median (IQR) | 33 | 39 (27.5-45) | 90 | 37.5 (25-49) | 0.8540 |
| Patients with positive Skin Prick Tests, n (%) | 33 | 23 (69.7) | 90 | 59 (65.6) | 0.8294 |
| Patients with positive Skin Prick Tests (perennial aeroallergens), n (%) | 33 | 13 (39.4) | 90 | 28 (31.1) | 0.3965 |
| Smoking status | 33 | 7 (21.2) | 90 | 31 (34.4) | 0.1906 |
| Smoking history, n (%) | 33 | 6 (18.2) | 90 | 25 (27.8) | 0.3520 |
| Current smoker, n (%) | 33 | 1 (3) | 90 | 6 (6.7) | 0.6733 |
| **Comorbidities** |  |  |  |  |  |
| Patients with anxiety/depression, n (%) | 33 | 1 (3) | 90 | 17 (18.9) | **0.0403** |
| Patients with GERD, n (%) | 33 | 4 (12.1) | 90 | 33 (36.7) | **0.0081** |
| Patients with bronchiectasis, n (%) | 33 | 3 (9) | 90 | 21 (23.3) | 0.1214 |
| Patients with atopic dermatitis, n (%) | 33 | 1 (3) | 90 | 3 (3.3) | 0.9999 |
| Patients with AERD, n (%) | 33 | 2 (6.1) | 90 | 7 (7.8) | 0.9999 |
| Patients with osteoporosis, n (%) | 33 | 2 (6) | 90 | 10 (11.1) | 0.5116 |
| Patients with CRSwNP, n (%) | 33 | 15 (45.5) | 90 | 38 (62) | 0.1073 |
| **Asthma outcomes** |  |  |  |  |  |
| Exacerbations / year, median (IQR) | 33 | 4 (2.3-8) | 90 | 5 (3-6) | 0.5506 |
| Patients who required ER / hospitalization, n (%) | 33 | 8 (24.2) | 90 | 26 (28.9) | 0.6569 |
| ACT, median (IQR) | 33 | 13 (8-16) | 90 | 12 (10-15) | 0.9285 |
| FEV_1_, %, median (IQR) | 33 | 68 (51.5-84.2) | 90 | 68.5 (57-86) | 0.5594 |
| FEV_1_, L, median (IQR) | 33 | 1.6 (1.2-2.2) | 90 | 1.7 (1.4-2.3) | 0.4889 |
| FVC, %, median (IQR) | 33 | 84 (66.5-95) | 90 | 86 (72-101) | 0.3857 |
| FEV_1_/FVC, %, median (IQR) | 33 | 73.8 (59.5-79) | 90 | 69 (58-77.1) | 0.2068 |
| FEF_25-75_, %, median (IQR) | 33 | 33 (22.2-49.3) | 90 | 32 (19-56) | 0.9127 |
| **Pharmacologic therapies** |  |  |  |  |  |
| High dose ICS-LABA, n (%) | 33 | 33 (100) | 90 | 90 (100) | 0.9999 |
| LAMA, n (%) | 33 | 20 (60.6) | 90 | 56 (62.2) | 0.9999 |
| Patients on OCS, n, (%) | 33 | 21 (63.6) | 90 | 60 (66.7) | 0.8309 |
| OCS, mg/die, median (IQR) | 33 | 7.8 (2.5-12.5) | 90 | 12.5 (2.5-25) | 0.2580 |
| Previous mAbs, n (%) | 33 | 6 (18.2) | 90 | 15 (16.6) | 0.7940 |
| **Biomarkers** |  |  |  |  |  |
| Blood eosinophils, cells/μL median (IQR) | 33 | 654 (390-790) | 90 | 500 (351-879) | 0.4753 |
| Blood basophils, cells/μL median (IQR) | 18 | 61 (32.8-91.5) | 57 | 48 (30-80) | 0.1533 |
| IgE, UI/ml, median (IQR) | 24 | 270 (90-376) | 80 | 181 (45-455) | 0.8296 |
| FeNO, ppb, median (IQR) | 24 | 47.5 (28.8-121) | 51 | 30 (15-59) | **0.0330** |

Data are presented as mean (SD), n (%) or median (interquartile range). *Abbreviations: BMI: body mass index, GERD: gastroesophageal reflux disease; AERD: aspirin-exacerbated respiratory disease; CRwNP: Chronic Rhinosinusitis with Nasal Polyps; ACT: Asthma Control Test; FEV_1_: forced expiratory volume in 1 s; FVC: forced vital capacity; FEF_25–75_%: forced expiratory flow 25–75%; ICS-LABA: inhaled corticosteroids- long-acting β-agonists; LAMA: long acting muscarinic agonists; OCS: oral corticosteroids; mAb: monoclonal antibody; IgE, immunoglobulin-E; FeNO: Fraction of Exhaled Nitric Oxide.* Bold entries highlight statistically significant *P*-values.

**Table E12.** Remission criteria: Zero exacerbations + zero OCS + ACT ≥20 + FEV_1_ decline <100mL from the best value of the first 12 months. Baseline characteristics comparing patients achieving sustained remission from month 12 to 24 vs non-remitters.

| **Zero exacerbations + zero OCS + ACT ≥20 + FEV_1_ decline <100mL from the best value of the first 12 months** | **n** | **Sustained remission** | **n** | **No remission** | ***P-value*** |
| --- | --- | --- | --- | --- | --- |
| **General characteristics** |  |  |  |  |  |
| Age, years, mean (SD) | 18 | 57.9 (10.5) | 105 | 57.8 (11.4) | 0.7786 |
| Female, n (%) | 18 | 11 (61.1) | 105 | 72 (68.6) | 0.1933 |
| BMI, mean (SD) | 18 | 28.5 (6.7) | 105 | 27.1 (5.3) | 0.7621 |
| Obese (BMI ≥ 30), n (%) | 18 | 4 (22.2) | 105 | 29 (27.6) | 0.7777 |
| Duration of disease, years, median (IQR) | 18 | 19 (10-25.3) | 105 | 18 (10-30) | 0.7905 |
| Age at onset, years, median (IQR) | 18 | 35.5 (25.3-45) | 105 | 40 (25-49) | 0.4952 |
| Patients with positive Skin Prick Tests, n (%) | 18 | 13 (72.22) | 105 | 69 (65.7) | 0.7877 |
| Patients with positive Skin Prick Tests (perennial aeroallergens), n (%) | 18 | 7 (38.9) | 105 | 34 (32.4) | 0.5971 |
| Smoking status | 18 | 4 (22.2) | 105 | 34 (32.4) | 0.5816 |
| Smoking history, n (%) | 18 | 4 (22.2) | 105 | 27 (25,7) | 0.9999 |
| Current smoker, n (%) | 18 | 0 (0) | 105 | 7 (6.7) | 0.5921 |
| **Comorbidities** |  |  |  |  |  |
| Patients with anxiety/depression, n (%) | 18 | 0 (0) | 105 | 18 (17.1) | 0.0717 |
| Patients with GERD, n (%) | 18 | 2 (11.1) | 105 | 35 (33.3) | 0.0923 |
| Patients with bronchiectasis, n (%) | 18 | 0 (0) | 105 | 24 (22.9) | **0.0224** |
| Patients with atopic dermatitis, n (%) | 18 | 0 (0) | 105 | 4 (3.8) | 0.9999 |
| Patients with AERD, n (%) | 18 | 0 (0) | 105 | 9 (8.6) | 0.3540 |
| Patients with osteoporosis, n (%) | 18 | 0 (0) | 105 | 12 (11.4) | 0.2109 |
| Patients with CRSwNP, n (%) | 18 | 9 (50) | 105 | 44 (41.9) | 0.6095 |
| **Asthma outcomes** |  |  |  |  |  |
| Exacerbations / year, median (IQR) | 18 | 6 (3-9) | 105 | 5 (3-6) | 0.6237 |
| Patients who required ER / hospitalization, n (%) | 18 | 5 (27.8) | 105 | 29 (27.6) | 0.9999 |
| ACT, median (IQR) | 18 | 11.5 (7.8-14.5) | 105 | 13 (10-15) | 0.1666 |
| FEV_1_, %, median (IQR) | 18 | 74.5 (49.8-88.3) | 105 | 68 (57-85.3) | 0.9840 |
| FEV_1_, L, median (IQR) | 18 | 1.7 (1.2-2.4) | 105 | 1.6 (1.3-2.2) | 0.8057 |
| FVC, %, median (IQR) | 18 | 84.4 (71.8.5-95.3) | 105 | 85 (68-101) | 0.7039 |
| FEV_1_/FVC, %, median (IQR) | 18 | 73.5 (59.5-79) | 105 | 69 (58-76.8) | 0.1522 |
| FEF_25-75_, %, median (IQR) | 18 | 33 (20.9-56.6) | 105 | 32 (19.5-51.8) | 0.6091 |
| **Pharmacologic therapies** |  |  |  |  |  |
| High dose ICS-LABA, n (%) | 18 | 18 (100) | 105 | 105 (100) | 0.9999 |
| LAMA, n (%) | 18 | 11 (61.1) | 105 | 71 (67.6) | 0.5971 |
| Patients on OCS, n, (%) | 18 | 13 (72.2) | 105 | 69 (65.7) | 0.7877 |
| OCS, mg/die, median (IQR) | 18 | 5 (2.5-12.5) | 105 | 12.5 (2.5-25) | 0.1828 |
| Previous mAbs, n (%) | 18 | 4 (22.2) | 105 | 17 (16.2) | 0.5085 |
| **Biomarkers** |  |  |  |  |  |
| Blood eosinophils, cells/μL median (IQR) | 18 | 680 (459-1022) | 105 | 500 (354-800) | 0.1087 |
| Blood basophils, cells/μL median (IQR) | 11 | 61 (47.8-108) | 64 | 50 (30-80) | 0.0919 |
| IgE, UI/ml, median (IQR) | 15 | 316 (125-457) | 89 | 179 (46-429) | 0.2041 |
| FeNO, ppb, median (IQR) | 14 | 66 (29-124) | 61 | 30.5 (15.8-56) | **0.0372** |

Data are presented as mean (SD), n (%) or median (interquartile range). *Abbreviations: BMI: body mass index, GERD: gastroesophageal reflux disease; AERD: aspirin-exacerbated respiratory disease; CRwNP: Chronic Rhinosinusitis with Nasal Polyps; ACT: Asthma Control Test; FEV_1_: forced expiratory volume in 1 s; FVC: forced vital capacity; FEF_25–75_%: forced expiratory flow 25–75%; ICS-LABA: inhaled corticosteroids- long-acting β-agonists; LAMA: long acting muscarinic agonists; OCS: oral corticosteroids; mAb: monoclonal antibody; IgE, immunoglobulin-E; FeNO: Fraction of Exhaled Nitric Oxide.* Bold entries highlight statistically significant *P*-values.

**Table E13.** Patients switched to different biologics during treatment.

| **Switch to:** | **n** | **12 months** | **n** | **24 months** |
| --- | --- | --- | --- | --- |
| Omalizumab (anti-IgE), n (%) | 232 | 2 (0.9) | 166 | 0 (0) |
| Benralizumab (anti-IL-5Rα), n (%) | 232 | 10 (4.3) | 166 | 15 (9) |
| Dupilumab (anti-IL-4Rα), n (%) | 232 | 2 (0.9) | 166 | 4 (2.4) |
| **Total, n (%)** | 232 | 14 (6) | 166 | 19 (11.4) |
